# The evolutionary landscape of host immunity genes involved in respiratory and other immune-related diseases, and their association with severe COVID-19 outcomes

**DOI:** 10.1101/2025.08.28.25334640

**Authors:** Christopher N. Cross, Alessandro Lisi, Faith C. Simmonds, Kareem Washington, Thomas Heinbockel, Michael C. Campbell

**Author notes:** To whom correspondence should be addressed **Corresponding author** E-mail address: Michael C. Campbell.

## Abstract

**Background:** Given its high mortality and broad societal impacts, the COVID-19 pandemic is a particularly notable global outbreak of a respiratory illness in the 21^st^ century. Although previous studies have identified several genes associated with COVID-19 susceptibility, relatively little is known about the genes contributing to severe COVID-19, including their evolutionary histories. In the current study, we analyzed *IL-4*, *TLR2*, *CCL2*, and *SLC11A1—*four immunity genes that have been implicated in severe COVID-19 and other immune-related diseases*—*in globally diverse populations from the 1000 Genomes Project. We also tested for associations between genetic variation in these genes and clinical COVID-19 phenotypes in more than 4,000 laboratory-confirmed COVID-19–positive individuals from Italy.

**Results:** Based on our analyses, we identified 72 single nucleotide polymorphisms (SNPs) across these genes as targets of positive selection, including several derived alleles shared with archaic Neanderthal and/or Denisovan genomes—a finding not previously reported in the literature. Furthermore, we found that common SNPs—implicated in respiratory diseases such as tuberculosis and chronic obstructive pulmonary disorder—were also under selection. Functional predictions based on *in silico* analyses revealed that a subset of selected alleles map to transcription factor binding sites and are predicted to affect binding affinity. In addition, our genetic association analyses uncovered significant correlations between derived alleles in the coding region of *TLR2* and COVID-19 severity. Interestingly, these candidate alleles occurred at relatively low frequency in western European and East Asian populations but were absent in populations of African and South Asian descent.

**Conclusions:** Overall, our study provides new insights into the evolution of biologically relevant immunity genes in the modern human lineage and highlights genetic variants that may underlie differential risk for severe COVID-19.

## Introduction

Novel coronavirus disease 2019 (COVID-19), caused by severe acute respiratory syndrome coronavirus 2 (SARS-CoV-2), was a public health crisis that resulted in more than 700 million positive cases and over 7 million deaths worldwide (https://www.worldometers.info/coronavirus/). While SARS-CoV-2 infection can lead to severe respiratory problems, other symptoms can include cardiovascular and neurological impairment [1–5]. Furthermore, data have shown that host genetics play a role in SARS-CoV-2 infection [6–11]. In particular, studies have identified host genes that encode receptors for coronavirus binding (*e.g.*, *ACE2*, *ANPEP*, *DPP4*) and proteases that facilitate entry of SARS-CoV-2 into cells (*e.g.*, *TMPRSS2*, *FURIN*, *TMPRSS11D*, *CTSL*, and *CTSB*), increasing COVID-19 susceptibility [7, 12, 13]. Genetic analyses have also demonstrated that alleles within these genes vary in frequency among populations [7, 13], potentially contributing to differential risk of infection in the absence of vaccination.

In addition, accumulating evidence has indicated that host genes contribute to severe COVID-19 after initial SARS-CoV-2 infection [12, 14–38]. For example, mRNA expression levels of Toll-like receptor genes, including *TLR2*, have been correlated with disease severity (commonly characterized by dysregulated inflammation and a cytokine storm) in SARS-CoV-2-positive patients [19, 39]; *in vivo* mice experiments further confirmed that the TLR2 protein was the innate sensor that triggered inflammatory cytokine expression in response to SARS-CoV-2 infection [19]. Prior analyses have also shown that cytokines, such as IL-4, IL-6, TNF-a, CXCL8, CXCL9, CXCL10, and CCL2, are present at elevated levels in the serum of COVID-19 patients with severe respiratory symptoms [40–44]. These findings suggest that COVID-19 severity may have a different genetic architecture involving cytokine and interleukin genes than COVID-19 susceptibility [45, 46]. In addition, multiple lines of evidence, including a meta-analysis of ∼400 host genetic susceptibility studies, have implicated a similar set of genes (specifically, *IL-4*, *TLR2*, *CCL2* and *SLC11A1*) in the onset of other respiratory and immune-related diseases, such as psoriasis, osteoarthritis, pulmonary tuberculosis (TB), inflammatory bowel syndrome (IBD), influenza, pneumonia, and severe acute respiratory syndrome (SARS) [47–63]. Finally, a recent analysis identified 13 genome-wide loci associated with lung, autoimmune, or inflammatory diseases that were significantly correlated with severe COVID-19 [6]. Thus, we hypothesize that genes known to play a role in respiratory and/or other immune-related illnesses, including their severity, may contribute to acute manifestations of COVID-19.

Despite the far-reaching consequences of COVID-19, the role of host genetics in severe disease has not been extensively examined. Furthermore, little is known about the evolutionary histories of immunity genes that have been implicated in severe COVID-19 and other immune-mediated diseases [56–58, 64, 65]. To overcome these gaps in knowledge, we examined *IL*-*4*, *TLR2*, *CCL2,* and *SLC11A1*—genes associated with an array of immune-mediated illnesses that also have been of long-standing interest to the authors—in 2,039 individuals from globally diverse populations in the 1000 Genomes Project to: 1) characterize patterns of nucleotide and haplotype variation within and between populations, and 2) infer the microevolutionary forces that have shaped these genetic patterns over time. In addition, we performed association analyses of clinical phenotypes and known mutations in our four genes implicated in respiratory and/or other immune-related illnesses in >4,000 laboratory-confirmed SARS-CoV-2-positive individuals enrolled in the GEN-COVID Multicenter Study in Italy. Overall, our study aims to characterize the evolutionary histories of key host immunity genes and provide new information regarding severe COVID-19 susceptibility.

## Results

### Patterns of Nucleotide Variation

We identified a total of 260 bi-allelic SNPs across the 8,690-base pair (bp) region of *IL-4* (**Figure 1**; **Table S1**). Of these 260 SNPs, 20 were found in exons, while the remaining 240 polymorphisms were present in non-coding regions (**Figure 1; Table S1**). Among the 20 exonic SNPs, nine were synonymous, 10 were nonsynonymous, and one was a stop gain mutation (*rs*550558636; **Table S1**). Interestingly, we found that nonsynonymous SNPs mainly occurred at very low frequency (< 1%) in all populations (**Table S1**). Similarly, most of the synonymous variants were either at low frequency (< 1%) or absent in populations, except for one SNP (*rs*2243251) which had a minor allele frequency (MAF) that ranged from 14% to 27%; the stop gain mutation was present only in the Mende of Sierra Leone with an MAF < 1% (**Table S1)**.

**Figure 1:**
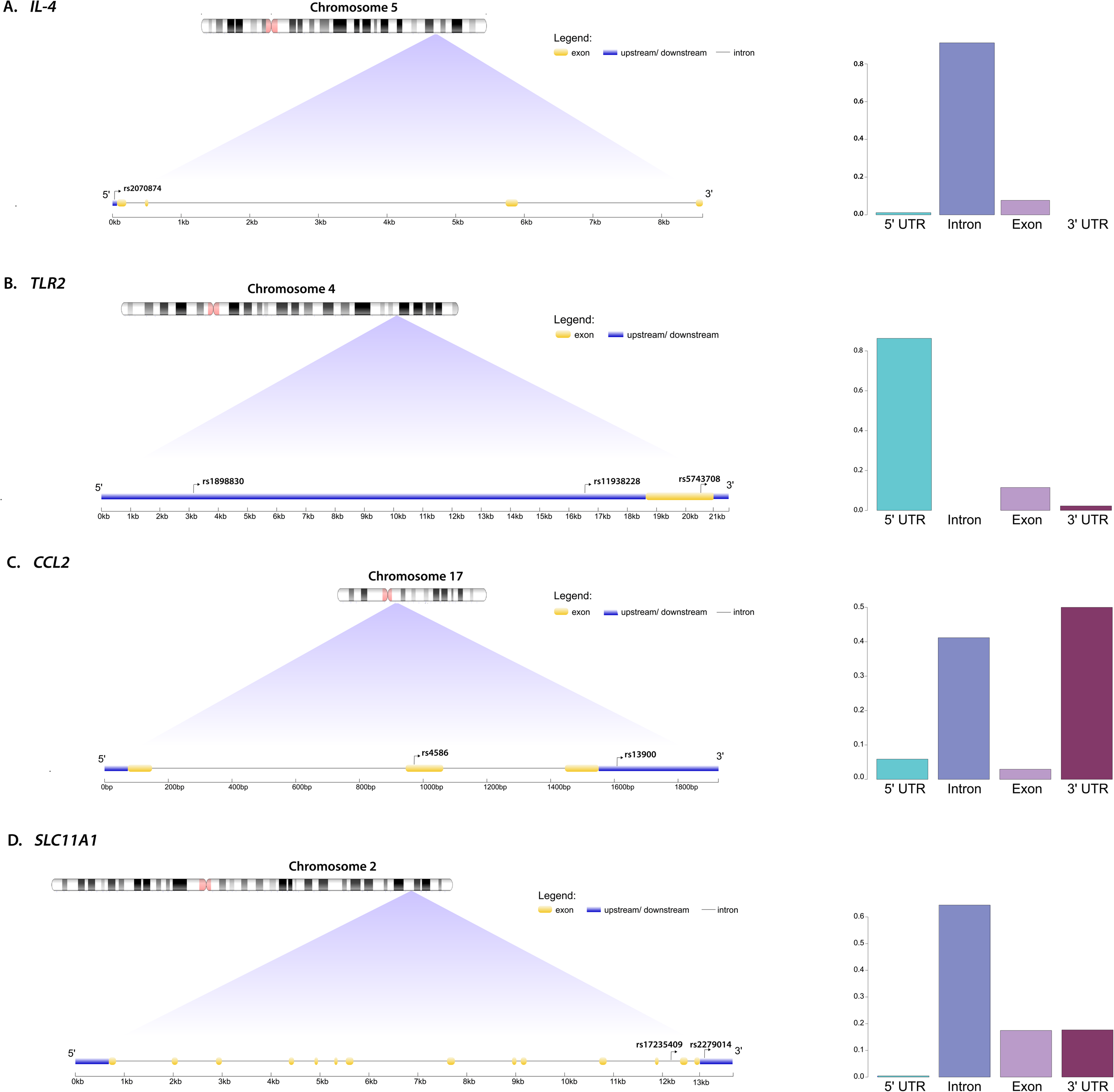
Gene structure and the distribution of polymorphisms across genic regions. This figure illustrates the chromosomal location and gene structure of *IL*-*4*, *TLR2*, *CCL2*, and *SLC11A1* along with the positions and *rs* identifiers of common SNPs previously associated with respiratory disease. Bar plots to the right of each gene diagram show the proportion of polymorphisms located in the 5′-untranslated region (5′-UTR), introns, exons, and the 3′-untranslated region (3′-UTR) of each immune-related gene (when applicable).

In the non-coding regions of *IL-4*, we identified five common SNPs (MAF ≥ 5%) in pooled populations (**Table S1)**; one of these SNPs (*rs*2070874) was located in the 5′-UTR, while the other four polymorphisms (*rs*2243281, *rs*734244, *rs*2243253, and *rs*2243261) were found in intronic regions. Furthermore, the MAF at these SNPs showed considerable variation across different populations. For example, the minor T-allele at *rs*2070874 exhibited frequencies ranging from 37.5% to 58% and from 73% to 86% in populations of African and East Asian descent, respectively (**Table S1)**. Prior studies have also linked the *rs*2070874 T-allele to severe respiratory illness and heightened immune responses [57, 66–68].

Across the 21,981-bp length of the intronless *TLR2* gene, we identified a total of 627 bi-allelic SNPs (**Figure 1**; **Table S2**). Within the single exon of this gene, we found 23 synonymous variants, 47 nonsynonymous variants, and two stop-gain variants. We also uncovered 541 and 14 polymorphisms in the 5′-UTR and 3′-UTR, respectively (**Table S2**). Furthermore, we identified 36 common SNPs in the 5′-UTR, 20 of which were found in all populations (none were present in the 3′-UTR). Interestingly, the MAF at these common sites differed widely among diverse populations (**Table S2**). For example, the minor T-allele at *rs*6811715 ranged from 33.9% to 49.2% frequency in populations of African descent but occurred at even higher frequency in non-Africans (between 47% and 53% in Europeans; between 50% and 60% in East Asians; **Table S2**); this allele was absent in South Asian populations. The minor T-alleles at *rs*13115230, *rs*11099894, *rs*7694512 also varied between 9% and 24% in populations of African descent but were observed at even higher frequency (≥ 29%) in non-African populations (**Table S2**). Similarly, the derived G-allele at *rs*1898830—which has been correlated with respiratory disease [69–71]—was present at relatively high frequency (from 29% to ∼60%) in non-African populations, particularly in Asia (**Table S2**). In contrast, the *rs*1898830 G-allele occurred at much lower frequency (from 7% to 23.3%) in African and African-descended populations (**Table S2**).

At *CCL2*, a total of 34 bi-allelic SNPs were identified across the 1,919-bp length of this gene (Figure 1). Of these bi-allelic SNPs, four were in exons, 11 in introns, two in the 5′-UTR, and 17 in the 3′-UTR (**Table S3**). Among the four exonic variants, two were synonymous (*rs*4586 and *rs*368861512) and two were nonsynonymous (*rs*148285031 and *rs*190996557). Furthermore, of the 34 SNPs that we identified, only four—*rs*13900 (3′-UTR), *rs*2857657 (intron), rs4586 (exon) and *rs*28730833 (intron)—had an MAF ≥ 5% (**Table S3**). In addition, the MAF of these common SNPs varied dramatically among populations. For example, the minor T-allele at *rs*13900 ranged from 20% to 28% frequency in populations of African descent while the same allele varied from 27% to 41% frequency in South Asians, from 29% to 36% in Europeans, and from 41% to 65% in East Asians (**Table S3**). Similarly, the rs4586 T-allele ranged from 13% to 37% frequency in African and African-descended populations but varied from 35% to 63% frequency in non-Africans (**Table S3**). Prior studies have also implicated *rs*13900 and rs4586 in gene expression regulation and/or respiratory disease susceptibility [65, 72, 73].

At *SLC11A1*, we identified 464 bi-allelic SNPs across the 14,622-bp length of this gene (**Figure 1**; **Table S4**). Of these SNPs, 75 were located in exons, 299 in introns, six in intronic splice regions, two in the 5′-UTR, and 82 in the 3′-UTR (**Figure 1; Table S4**). We also observed eight common SNPs in exons, 41 in introns, and eight in the 3′-UTR (**Table S4)**. Furthermore, the MAF at common exonic SNPs differed considerably between African and non-African populations. For example, the minor *rs*17221959 T-allele occurred at a frequency no greater than 15.5% in populations of African descent, whereas this allele varied between 28.3% and 40.3% frequency in non-Africans (**Table S4)**. Conversely, the minor *rs*7576974 T-allele spanned from 0.08% to 26% frequency in populations of African descent but occurred at <0.02% frequency in non-Africans (**Table S4)**.

We also noted frequency differences at common intronic variants at *SLC11A1* across populations. For example, the minor *rs*2290708 T-allele varied between 19% to 35% frequency in Europeans but was present at <17.9% frequency in other global populations (**Table S4**). Likewise, the T-alleles at *rs*13011285 and *rs*17228995 ranged from 20.7% to 34.1% in European populations but occurred at lower frequency in populations of African, South Asian, and East Asian descent (**Table S4**). Furthermore, the derived *rs*2279014 T-allele in the 3’-UTR—associated with decreased risk for *Mycobacterium avium* complex infection [74]—was found at ∼40% frequency or higher in most populations, except in East Asians where the frequency of this allele varied between 18.8% and 30% (**Table S4**).

### Tests of Neutrality

To investigate if patterns of nucleotide variation evolved neutrally, we calculated Fay and Wu’s *H* (*H*) and Tajima’s D (*D*_T_) statistics for *IL*-*4*, *TLR2*, *CCL2* and *SLC11A1* in each population with DnaSP [75] (**Tables S5; Tables S6; Tables S7; Tables S8**). We also simulated the expected *H* and *D*_T_ values under different models of population growth to assess the statistical significance of observed values (**Tables S9-S16**). Using this approach, we found a mix of positive and negative *H* values at *IL-4* in non-African populations with significant negative *H* statistics observed in East Asians. In contrast, populations of African descent generally exhibited positive *H* values (**Table S9**). At *TLR2*, *H* statistics were consistently negative in non-Africans with significant values occurring in European, South Asian, and East Asian populations. We also observed a pattern of significantly negative *H* statistics at *TLR2* in populations of African descent (**Table S10**). Our analyses of *CCL2* revealed a pattern of negative *H* statistics in non-African populations (**Table S11**). We observed a similar trend in Africans with significant values occurring in the East African Luhya and West African Yoruba. At *SLC11A1*, significantly negative *H* values were found in both African and non-African populations (**Table S12**).

Finally, although the *D*_T_ statistics were a mix of negative and positive values at *IL-4*, *TLR2*, and *CCL2* in both Africans and non-Africans, these values were generally more positive than expected under different population growth models (**Tables S13-S15**). By contrast, at *SLC11A1*, we observed significantly negative *D*_T_ statistics in populations of African descent and consistently negative *D*_T_ statistics in non-Africans (though these values were not significant; **Table S16**).

### Long-range haplotype homozygosity

We calculated *i*HS and *nS*_L_ statistics for each population to examine patterns of haplotype homozygosity surrounding the *IL-4*, *CCL2*, *TLR2*, and *SLC11A1* genes using the selscan and Polaris packages [76, 77]. Based on these analyses, we found outlier | *nS*_L_ | statistics at several *IL-4* variants— specifically, *rs*199693275, *rs*734244, *rs*58349986, *rs*56408050 and *rs*56174361—primarily in non-African populations (**Table 1; Table S17; Figures S1-S2**). Furthermore, we observed short extended haplotype homozygosity (EHH) around *rs*58349986 and *rs*56174361 in non-Africans, particularly in Europe and Asia. At *TLR2*, we identified SNPs with extreme | *i*HS | and/or | *nS*_L_ | statistics in African and non-African populations (**Table 1**; **Tables S18-S19; Figures S4-S5**). In several cases, we also detected unusually long haplotypes around either the derived or ancestral alleles at these loci. (**Figure S6**). Most notably, we observed long-range EHH surrounding the derived *rs*7694512 T-allele and derived *rs*11099894 T-allele, spanning up to 700,000 kbs, in African and South Asian populations (**Figure S6**). Additionally, the derived G-allele at *rs*1898830—which has been previously correlated with a decrease risk for TB development [71]—was found on an extended haplotype background in these populations (**Figure S6**).

**Table 1.**
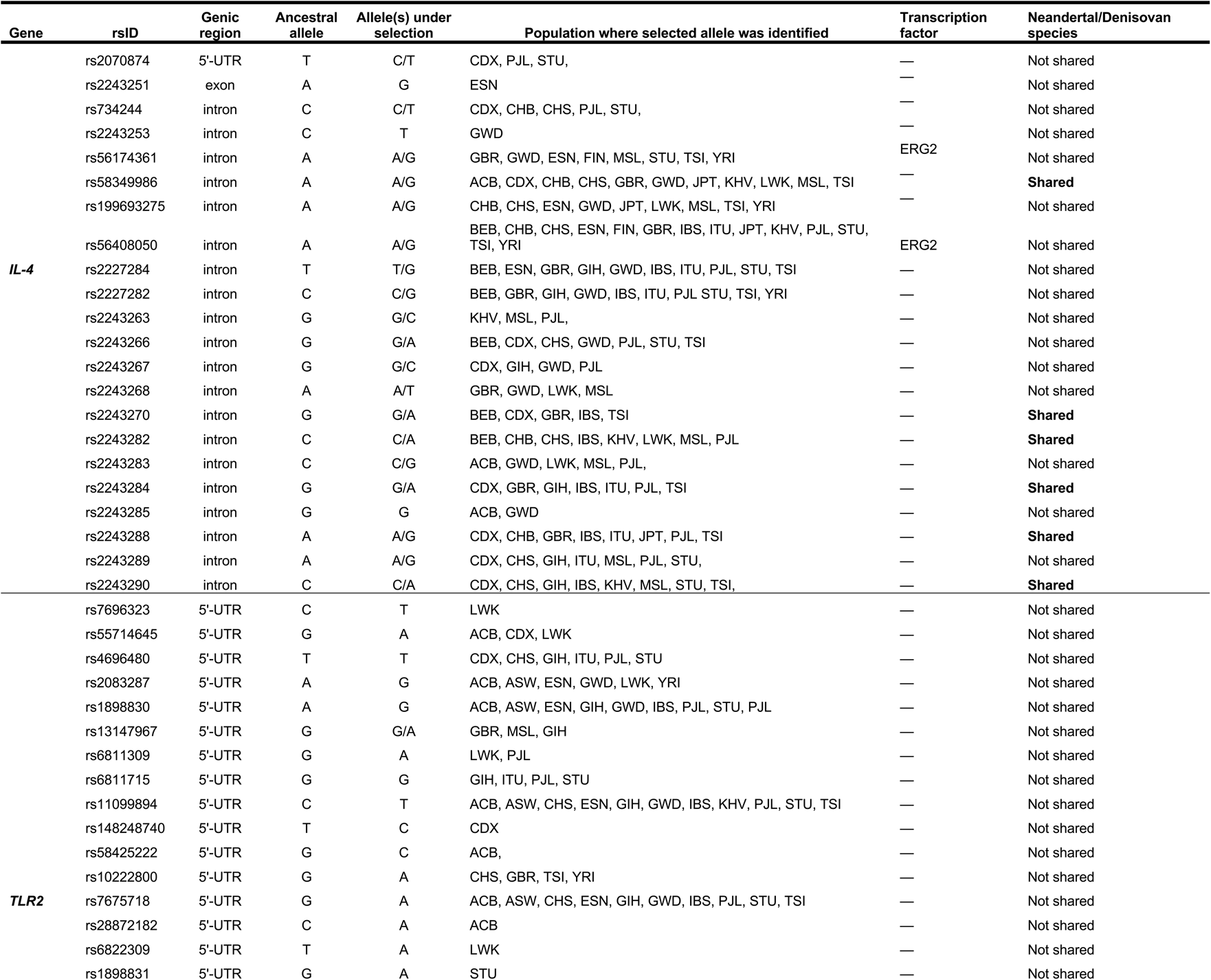
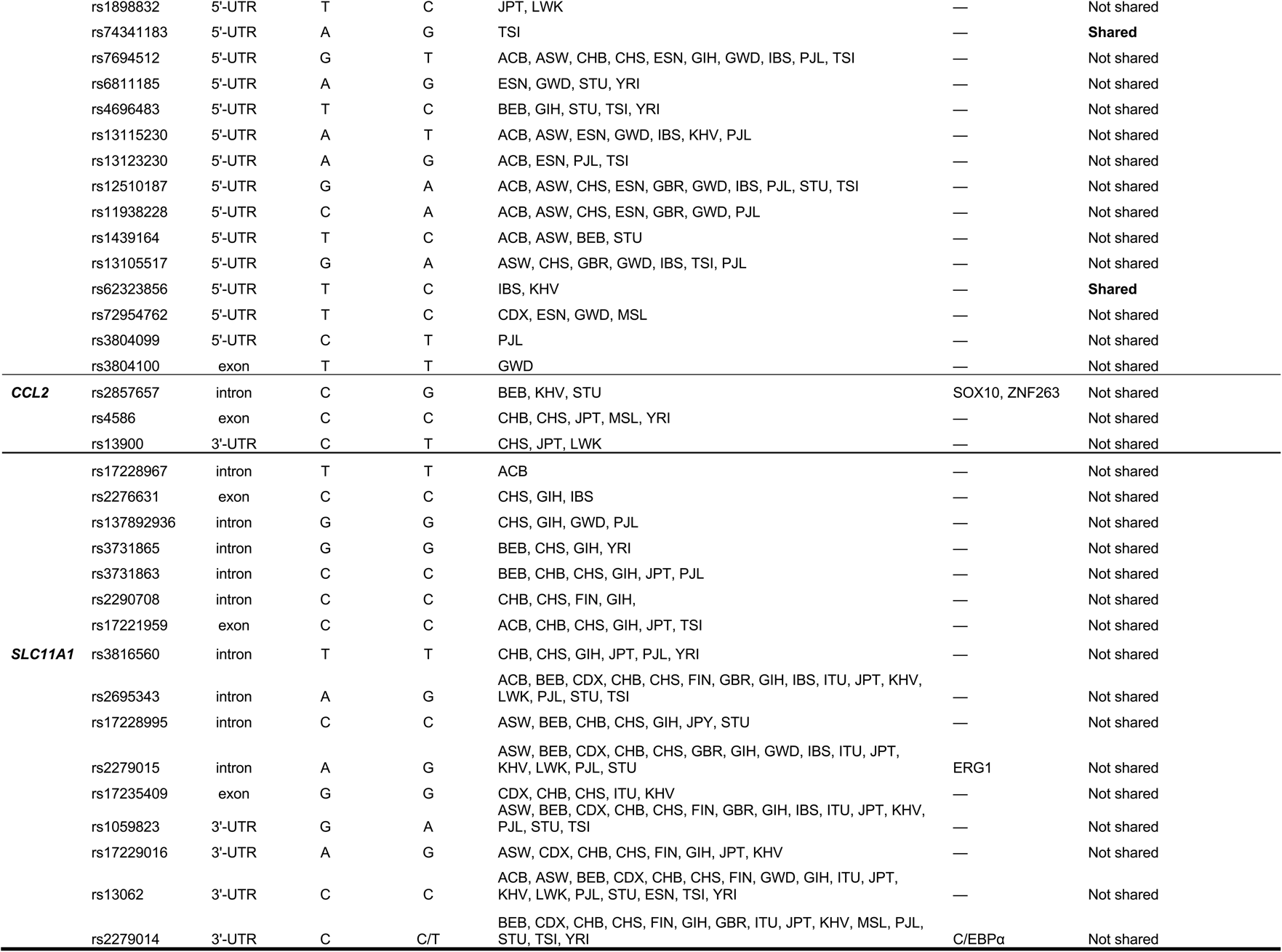
Inferred targets of selection and their predicted functional impact. The SNPs identified as adaptive loci are given in the Table. These loci were inferred to be targets of selection based on | *i*HS |, | *nS*_L_ |, EHH, and CLUES2 results. For each of these selected sites, we listed the rs identifier, the genic region in which SNPs are located, the ancestral allele, the allele believed to be the target(s) of selection, the population in which the selected allele(s) was found, and the known transcription factor that binds to the transcription factor binding site (TFBS) where the SNP occurred; “—” means that a given SNP was not found in a known TFBS according to the SNP2TFBS database (last accessed 08/14/2025). The full population names for the abbreviations in the Table can be found in the Methods section.

At *CCL2*, we identified two SNPs (*rs*2857657 and *rs*4586) that exhibited extreme | *i*HS | or | *nS*_L_ |statistics in global populations (**Tables S20-S21; Figures S7-S8**). Specifically, *rs*2857657 had an extreme positive *i*HS statistic, indicating selection on the derived G-allele, in the Afro-Caribbean and Iberian populations (**Tables S20-S21; Figures S7-S8**), while *rs*4586 exhibited negative outlier *i*HS statistics favoring the ancestral C-allele in populations of African, European, and East Asian descent (**Tables S20-S21**). As with *TLR2*, we plotted the decay of haplotype homozygosity from these core SNPs using the EHH statistic [78]. This analysis revealed unusually long haplotype structure—spanning ∼200 kbs around the derived *rs*2857657 G-allele—in the Afro-Caribbean and Iberian populations (**Figure S9**). Furthermore, we detected extensive EHH on chromosomes carrying the ancestral rs4586 C-allele (relative to chromosomes with the derived allele at the same site) in East Asian populations (**Figure S9**).

Finally, we observed distinct patterns of haplotype homozygosity around the *SLC11A1* gene when comparing African and non-African populations. Notably, several SNPs within *SLC11A1* exhibited outlier | iHS | and/or | *nS*_L_ | statistics in South Asian, European, and East Asian populations (**Tables S22 and S23; Figures S10 and S11**). We further examined the decay of haplotype homozygosity around these outlier SNPs using the EHH statistic (**Figure S12**). Based on this analysis, we found extended haplotype homozygosity (EHH) around the ancestral C-allele of rs2279014 in South Asian, European, and East Asian groups (**Figure 2; Figure S12**). Similarly, we detected unusually long haplotypes around the derived rs2279015 G-allele in South and East Asians (**Figure 3; Figure S12**). These variants are of particular interest as both are predicted to affect transcription factor binding. Furthermore, the rs2279014 T-allele is linked to decreased susceptibility to *Mycobacterium avium* complex infection [74]. Comparatively, African and African-descended populations did not show outlier | *i*HS | and | *nS*_L_ | values at these loci.

**Figure 2:**
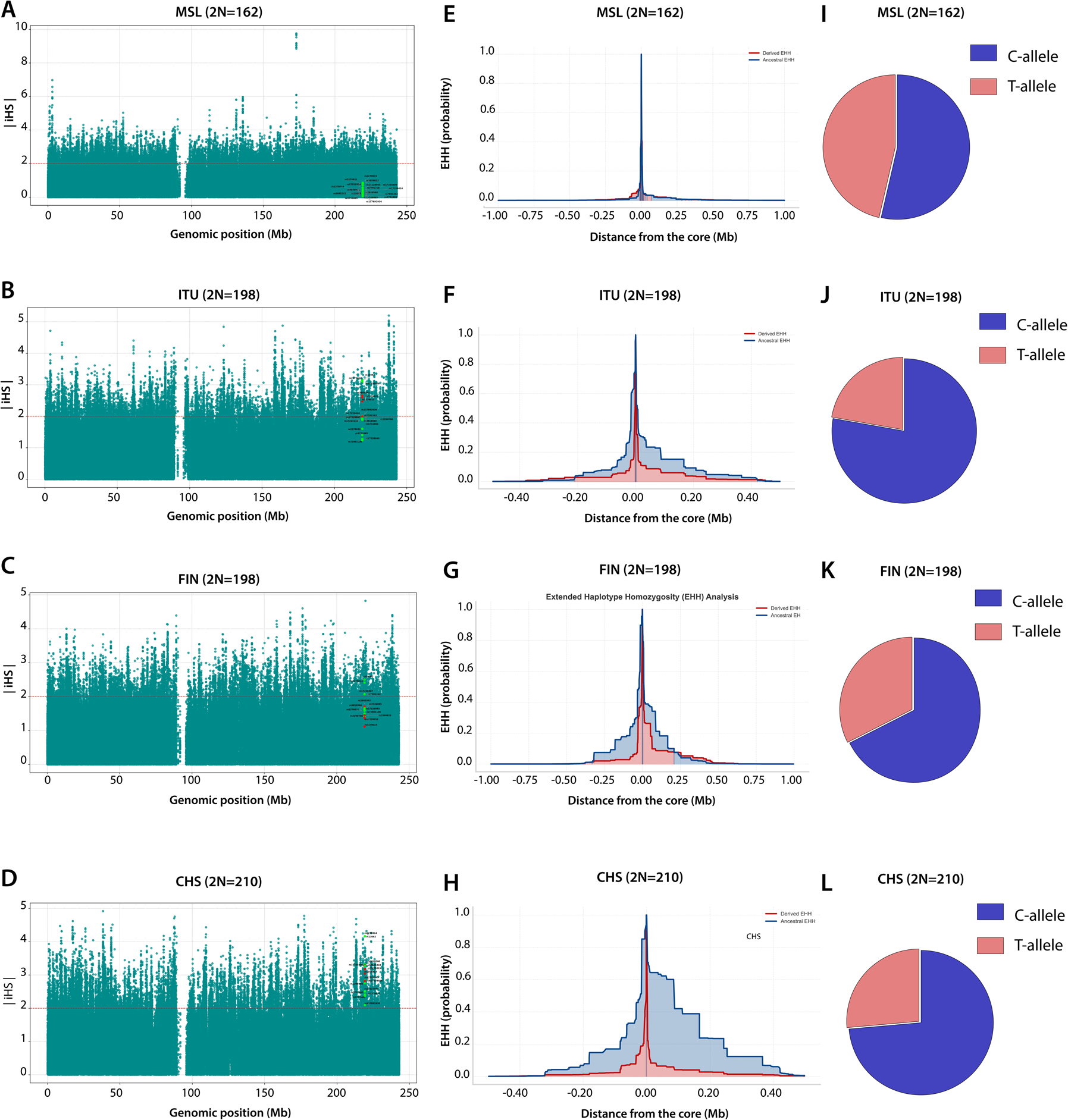
Signatures of positive selection and allele frequencies patterns at rs2279014 in a subset of global populations. Panels A-D show Manhattan plots of standardized | *i*HS | statistics on Chromosome 2 in exemplar populations from the 1000 Genomes Project. The horizontal dashed line indicates the threshold for identifying outlier *i*HS statistics. Red dots indicate the derived allele at SNP loci while the green dots denote ancestral alleles at SNPs across *SLC11A1*; we also listed their corresponding rs identifiers. Panels E-H show the decay of extended haplotype homozygosity (EHH) surrounding the functional *rs*2279014 SNP. The x-axis is the distance in megabases (Mb) upstream (negative values) and downstream (positive values) from the *rs*2279014 core site on the forward strand; the *y*-axis is the probability that two chromosomes are homozygous at all SNPs for the interval from the core site to distance *x*. Blue and red shading indicates the decay of homozygosity on chromosomes carrying the ancestral and derived alleles, respectively, from the core site. The frequencies of the ancestral and derived alleles at *rs*2279014 are given for each population. The additional *i*HS and EHH plots for other populations are provided in the Supplementary Materials.

**Figure 3:**
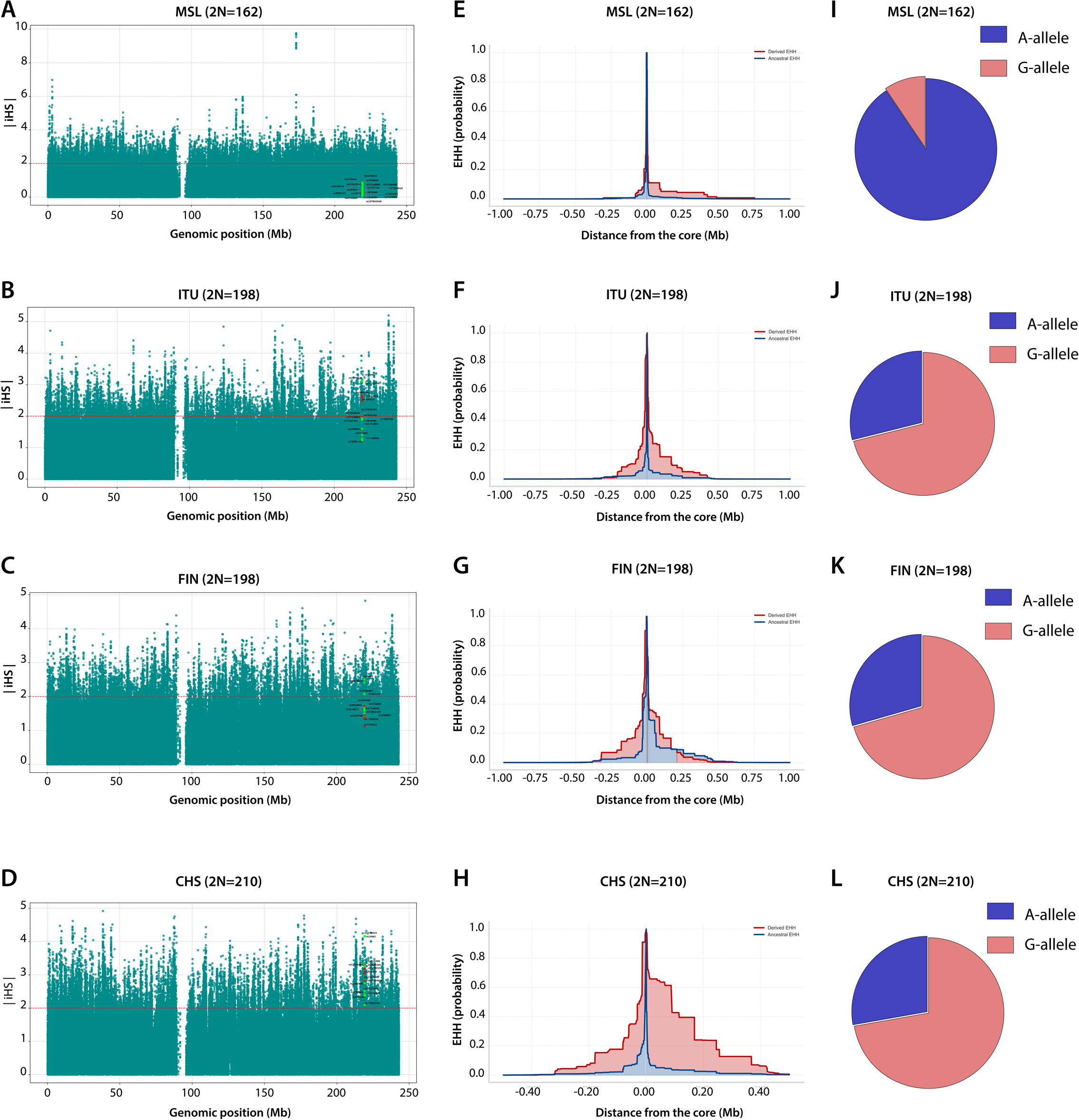
Signatures of positive selection and allele frequency patterns at rs2279015 in a subset of populations. Panels A-D illustrate Manhattan plots of standardized *i*HS statistics on Chromosome 2 in exemplar populations from the 1000 Genomes Project. The horizontal dashed lines represent the cut-off for extreme | *i*HS | scores (| *i*HS | = 2). Red dots denote the derived alleles at *SLC11A1* SNPs while the green dots indicate ancestral alleles at SNPs in the same gene. Panels E-H display EHH around the functional *rs*2279015 SNP (core site) in exemplar populations. Specifically, these plots show the decay of identity of haplotypes on chromosomes carrying the derived allele (red shading) versus the ancestral allele (blue shading) at *rs*2279015 with increasing distance from the core site. The x-axis shows the distance of homozygosity from the core SNP (at position zero), with negative numbers indicating upstream regions and positive values signifying downstream regions on the forward strand. The EHH probabilities are given on the y-axis. The frequencies of the ancestral and derived alleles at *rs*2279015 are provided for each population. Additional *i*HS and EHH plots for other populations are available in the Supplementary Materials.

### Selection coefficient estimates

As a complementary analysis, we applied the CLUES2 algorithm to *IL-4*, *TLR2*, *CCL2*, and *SLC11A1* in each population at different time depths (i.e., 1,000, 500, and 200 generations ago) to assess temporal shifts in selection intensity [79–81]. Our results indicated that selection pressure on SNPs in *IL-4* likely began at a deep point in time. For example, we observed significant *s* estimates for derived alleles at rs56174361 in African populations at 1000 generations ago (corresponding to 28,000 ya; **Figure 4; Table S24**). Furthermore, we detected significant *s* for ancestral alleles at *rs*2227282 and *rs*2227284 in African populations (**Figure 4; Table S24**). In contrast, selection appeared to target the ancestral allele at *rs*56174361, as well as the derived alleles at *rs*2227282 and *rs*2227284 in different populations (**Figure 4; Table S24**). Interestingly, this pattern of selection on derived and ancestral alleles at the same polymorphic site occurred across *IL-4*; in many cases, the inferred timing of these selection events dated to at least 1000 generations ago, suggesting deep and complex selection histories at this gene.

**Figure 4:**
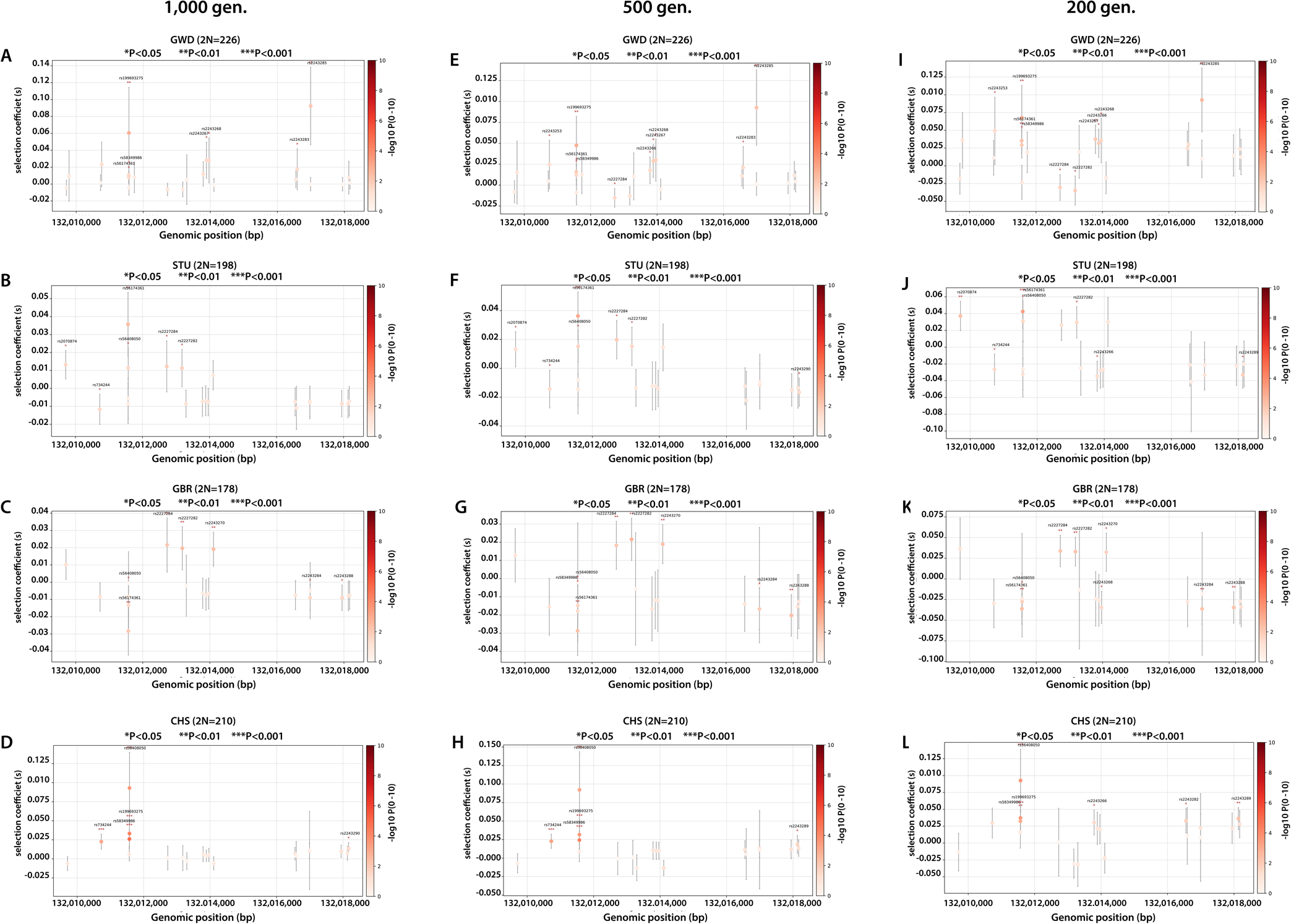
Estimated selection coefficients for SNP loci in *IL-4* in global populations. Panels A-D, Panels E-H, and Panels I-L show *s* estimates for SNPs in a subset of global populations together with the rs identifiers for SNPs that exhibit significant s values (*P* < 0.05) across *IL-4* at three different time depths (1,000 generations ago, 500 generations ago, and 200 generations ago, respectively). Each dot represents an estimated *s* statistic, and the vertical bars indicate the confidence intervals around the estimate. Negative *s* estimates indicate selection on ancestral alleles while positive *s* values signify selection on derived alleles. The color of each dot indicates the level of significance associated with the *s* statistics in addition to the asterisks.

Similarly, at *TLR2*, we found evidence for strong selection favoring derived alleles at 1,000 generations ago (28,000 ya) in Africa. For example, in the Yoruba population from Nigeria, *s* estimates for *rs*2083287 and *rs*6811185 were 0.2942 (CI: 0.00664–0.0522) and 0.01293 (CI: −0.00166–0.02753), respectively (**Table S25**), indicating selection on derived alleles at these sites; however, we did not observe extensive EHH around these polymorphic sites. In contrast, we did observe non-significant *s* estimates for derived alleles at other SNPs (such as *rs*12510167, *rs*7694512, *rs*11099874, *rs*7694512, and *rs*1311523), which exhibited extensive EHH in populations of Africa descent. This finding suggests that strong selection may have occurred more recently. One notable exception to this pattern was *rs*13123230 which showed long-range EHH and a significant *s* estimate at 1,000 generations ago (28,000 ya) in the Esan from Nigeria (**Table S25; Figure S6**). However, upon further examination, we found that *s* was 0.01405 (CI: 0.00612– 0.02199) at 1,000 generations ago, decreased to 0.00386 (CI: −0.17166–0.17938) at 500 generations ago (14,000 ya), and then dramatically increased to 0.01683 (CI: −0.26538–0.29903) at 200 generations ago (5,600 ya; **Table S25**), indicating a more recent increase in selection pressure. Similarly, outside of Africa, we observed progressively stronger *s* estimates from 28,000 ya to 5,600 ya at multiple SNPs in populations, including sites such as rs62323856 and rs74341183 which are polymorphic in the Neandertal and/or Denisovan genomes (**Table S25**).

At *CCL2*, we found that the strength of selection at loci in this gene differed among populations (**Table S26**). For instance, we observed a significant *s* estimate for the ancestral C-allele at rs4586 in Africans (Mende and Yoruba) and East Asians (Japanese) but not in other populations (**Table S26**). We also found significant *s* estimates at rs13900 favoring the derived allele in Africans (Luhya) and East Asians (Han Chinese and Japanese), as well as significant *s* for *rs*2857657 in the Tuscan Italian population (**Table S26**).

At *SLC11A1*, we detected significant *s* statistics at several polymorphic sites in African and European populations at least 28,000 ya (**Table S27**). This deeper timeframe of selection is further corroborated by the absent of extensive EHH in these populations [78]. In contrast, we observed instances of strong selection at sites at a more recent time depth in South Asian and East Asian populations (**Table S27**). For example, rs2695343 exhibited significant *s* estimates favoring the derived allele at least 200 generations ago (5,600 ya). Intriguingly, we also detected significant *s* estimates for derived allele at rs2279015 and rs1059823 that progressively increased over time (from 28,000 ya to 5,600 ya) in South and East Asian populations (**Table S27**), which likely contributed to the observed long-range EHH in these groups (**Table S27**). Finally, unlike the previous three genes, we found very few instances in which both ancestral and derived alleles at the same site were targeted by selection in different populations (**Table S27**).

### Population differentiation

We assessed among-population differentiation by calculating *F*_ST_ statistics at polymorphic sites within our genes. To identity *F*_ST_ outliers, we compared observed values to an empirical distribution consisting of *F*_ST_ estimates calculated at ∼8.1 million SNP randomly chosen across the genome. Based on this approach, we identified nine SNPs at *IL-4* (*rs*2243288, *rs*2243289, *rs*2243290, *rs*2243284, *rs*2243282, *rs*2243270, *rs*561556119, rs2227282, *rs*2227284) that had outlier *F*_ST_ statistics (**Table S1**). Notably, the derived alleles at these sites (specifically, *rs*2243288, rs2243290, rs2243284, rs2243282) were also present in the Neandertal and/or Denisovan genomes (**Table S1**). At *TLR2*, we detected outlier *F*_ST_ statistics at *rs*1439164, *rs*6811185, *rs*2083287, *rs*1898831, and *rs*4315760 (**Table S2**). We also identified a single outlier *F*_ST_ estimate at *rs*2279015 in the *SLC11A1* gene (**Table S4**); however, we did not detect extreme *F*_ST_ values at loci in *CCL2* (**Table S3**). Interestingly, the majority of the highly differentiated SNPs listed above, including those sites of archaic origin, were inferred to be under selection in contemporary populations based on our *i*HS, *nS*_L_, EHH, and/or CLUES2 analyses (**Tables S17-S23; Figures S1-S12**).

### Distribution of haplotype variation

At *IL-4*, we inferred 296 distinct haplotypes, nine of which were common (H44, H74, H85, H103, H115, H197, H234, H241, and H287) in populations of African descent (**Tables S28-S30**). Together, these common haplotypes comprised 51% to 58% of all haplotypes in these populations (**Tables S28-S30**). In contrast, eight common haplotypes (H44, H69, H82, H176, H197, H198, H234, and H241) comprised ∼80% to 93% of all haplotypes in non-African populations (**Tables S28-S30**). Further analyses also revealed that a subset of these common haplotypes (H44, H69, H82, H176, and H234) accounted for more than 78% of all haplotypes in Europeans and South Asians; these same haplotypes only constituted between 47.4% and 74.6% of the total haplotypes in East Asian populations (**Tables S28-S30**).

At *TLR2*, we extracted a total of 599 distinct haplotypes in pooled global populations (**Tables S31-S33)**; eight common haplotypes (H11, H30, H231, H265, H361, H402, H438, and H580) comprised ∼38% to 44% of the variation present in African and African-descended populations (**Tables S31-S33**). In comparison, 12 common haplotypes (H2, H40, H65, H96, H98, H108, H227, H231, H482, H507, H532, and H540) constituted ∼49% to 80% of the haplotypes in non-Africans (**Tables S31-S33)**. Our analyses also showed that a few of these haplotypes largely contributed to the variation in these populations. For example, H231 accounted for ∼40% to 46% of all the haplotypes in South Asians, while H231 comprised ∼24% to 25% and ∼24% to 32.9% of the haplotypes in Europeans and East Asians, respectively (**Tables S31-S33**). In contrast, the maximum frequency of haplotypes was 15% in populations of African descent (**Tables S31-S33)**.

At *CCL2*, we identified 35 distinct haplotypes in pooled global populations (**Tables S34-S36)**, five of which were common (H2, H9, H11, H19, and H29). Our results also showed that H9, H11, H19, and H29 constituted 92%-97% and 97% to 100% of the total variation in African and non-African populations, respectively (**Tables S34-S36**), indicating a strong skew toward a small set of dominant haplotypes at this gene.

At *SLC11A1*, we found a total of 619 distinct haplotypes in pooled global populations (**Tables S37-S39**); 11 common haplotypes (H102, H159, H179, H204, H215, H254, H269, H334, H445, H484, and H491) comprised between ∼28% and 54% of all haplotypes in populations of African descent (**Tables S37-S39**). In contrast, eight common haplotypes (H150, H223, H284, H321, H334, H340, H484, and H491) comprised between ∼50% and 75% of all haplotypes in non-Africans (**Tables S37-S39**). Notably, only two common haplotypes (specifically. H334 and H484) were shared between African and non-African populations (**Tables S37-S39**), revealing a high degree of divergence between these broad groups at this gene.

### Inferred haplotype relationships

We reconstructed the evolutionary relationships among haplotypes at each gene using the median joining network method [82](**Figure 5**). This analysis revealed a divergent geographic distribution of several common *IL*-*4* haplotypes (**Figure 5; Table S40**). Specifically, H44, H82, and H69 were found mainly in European and South Asian populations, while H234 and H197 were more prevalent in East Asians (**Figure 5**). We also observed African-specific haplotypes (i.e., H74, H85, H103, H115, and H287) at low to moderate frequency; the presence of reticulations among these African haplotypes suggests historical recombination (**Figure 5**)[83]. Our previous analyses also showed that both ancestral and derived alleles at the same SNP (e.g., *rs*2243288, *rs*2243290, *rs*2243284, and *rs*2243282) were subject to positive selection in different populations. Many of the ancestral alleles at these sites reside on haplotypes H44, H82, and H69 which are common in European and South Asian populations (**Figure 5**). Conversely, many of the derived alleles at the same SNPs were found on a different set of haplotypes (H176, H197, H234, and H241) that were prevalent in East Asians (**Figure 5**).

**Figure 5:**
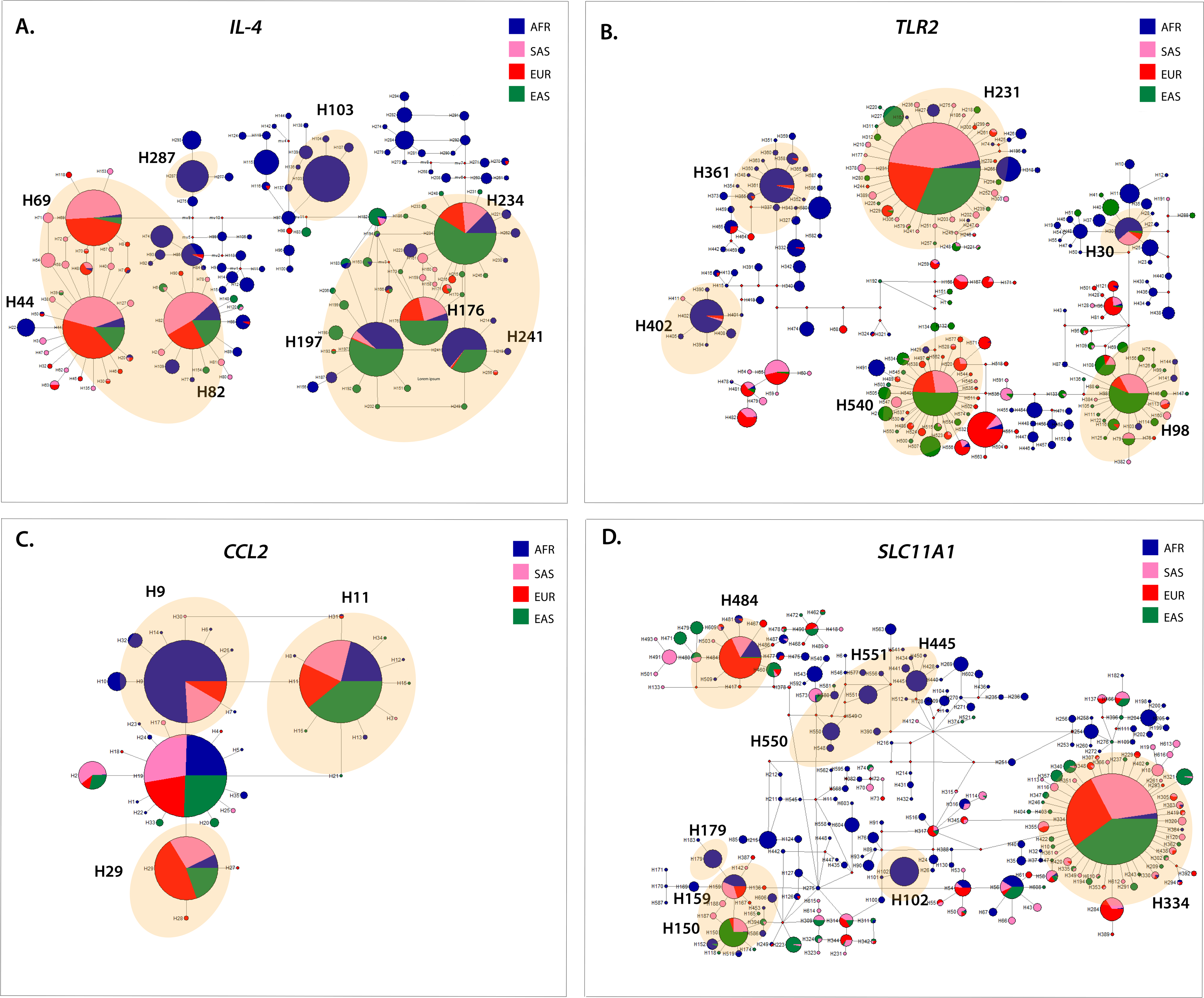
Haplotype genealogies of global variation across *IL*-*4*, *TLR2*, *CCL2*, and *SLC11A1*. Haplotype relationships were reconstructed using the median-joining algorithm implemented in Network 5.0 [82]. The resulting phylogeny consists of haplotypes (or nodes), and the size of each haplotype is proportional to the number of chromosomes with that given haplotype. The colors within the nodes (or haplotypes) indicate the frequency of that haplotype in continental populations. Specifically, blue represents African and African-descended populations (AFR); pink signifies South Asian populations (SAS); red denotes European populations (EUR); green indicates East Asian populations (EAS). The lines connect the nodes in the shortest possible tree. High-frequency haplotypes carrying alleles inferred to be under selection are shaded in orange, and the corresponding haplotype identifiers (e.g., H69, H98) are listed near them. The genomic coordinates and rs identifiers for selected SNPs on these common haplotypes are given in Table S40 (*IL-4*), Table S41 (*TLR2*), Table S42 (*CCL2*), and Table S43 (*SLC11A1*).

At *TLR2*, H98, H231 and H540 were the most frequent haplotypes in the phylogeny and were surrounded by low-frequency haplotypes in a “star-like” pattern, suggestive of a recent expansion of variation (**Figure 5**). Among these haplotypes, H231 contained the largest number of selected alleles compared to H98 and H540, and H231 had the highest frequency in the network (**Figure 5**). Furthermore, H98 and H540 showed a striking geographic distribution. In particular, haplotypes H98 and H540 were present only in non-Africans while haplotypes H265, H361, H402 were found mainly in populations of African descent (**Figure 5**; **Table S41**). Our analyses also revealed that different haplotypes carried distinct combinations of selected alleles. For example, the derived alleles at *rs*13147967 and *rs*1898831 occurred on several haplotypes (e.g. H98, H231, and H540) that were widespread in Eurasian populations, while the derived alleles at *rs*2083287 and *rs*6811185 existed on haplotypes (e.g., H361, H402) that were more common in populations of African descent (**Figure 5; Table S41**).

At *CCL2*, H9, H11, H19, and H29 haplotypes occurred at high frequency in global populations, except for H9 which was absent in East Asia (**Figure 5; Tables S37-S39)**. Notably, H9 carried the ancestral *rs*4589 C-allele, which has been inferred to be under strong selection in Africans. Haplotype H11 contained both the *rs*4589 C-allele and the derived *rs*13900 T-allele, which have significant *s* estimates in East Asian populations (**Table S42**). Comparatively, H29 carried the derived *rs*2857657 G-allele and the derived *rs*13900 T-allele; the highest frequency of this haplotype occurred in Europe (**Figure 5)**.

Finally, at *SLC11A1*, the common H334 and H484 haplotypes were found primarily in non-African populations (**Figure 5; Table S43**). Specifically, H334 occurred at low frequency (∼2%) in populations of African descent (**Table S26; Table S27**), while it was present at a much higher frequency in non-Africans (i.e., ∼40%, ∼29% and ∼29% in East Asians, Europeans, and South Asians, respectively; **Tables S37-S39**). H484 was observed at relatively high frequency in Europeans (∼60%) but at much lower frequency in populations of South Asian and African descent (∼20%; **Tables S37-S39**); H484 was absent in East Asian populations. Furthermore, H334 and H484 harbored multiple alleles with significant *s* estimates (**Figure 5**; **Tables S37-S39**). Likewise, haplotypes specific to populations of African descent (such as H454 and H551) carried alleles with significant *s* values (**Table S43**). These African-specific haplotypes also exhibited reticulations—represented by loops in the haplotype network—which indicate past recombination (**Figure 5**).

### *In silico* functional analysis

Given that gene expression differences are known to contribute to disease susceptibility and/or severity [84–86], we analyzed the common SNPs that were inferred to be adaptive using the SNP2TFBS tool [87], which predicts whether or not polymorphisms of interest occur in transcription factor binding sites (TFBSs) [87, 88]. This analysis identified two SNPs at *IL-4* (rs56174361 and rs56408050) which were inferred to affect the binding of the ERG2 transcription factor (TF). Furthermore, one SNP at *CCL2* (rs2857657)—located in a TFBS—is predicted to affect the binding of the SOX10 and ZNF263 transcription factors (**Table 1**). At *SLC11A1*, *rs*2279014 and *rs*2279015 are predicted to alter the binding of the CEBPA and ERG1, respectively (**Table 1**). In contrast, none of the SNPs at *TLR2* overlapped with known TFBSs in the SNP2TFBS database.

### Genotype-phenotype analysis

Because we hypothesized that loci associated with immune-mediated diseases could also influence severe COVID-19 outcomes, we focused our analyses on previously identified candidate loci associated with immune-related illnesses in laboratory-confirmed SARS-CoV-2-postitive individuals in two GEN-COVID cohorts. First, we examined comorbidities known to increase the risk of severe COVID-19 along with genotypes from our four genes in a cohort of 1,141 SARS-CoV-2-positive individuals. Using a linear mixed model approach, we detected a significant association between the missense *rs*139227237 mutation in the coding region of *TLR2* and organ transplant status (P < 0.03; **Table S44**). Notably, the *rs*139227237 C-allele [p.(Phe217Ser)] was present in SARS-CoV-2-infected individuals who received an organ transplant and absent in SARS-CoV-2-positive individuals without a transplant history. This finding is particularly relevant given that organ transplant recipients are known to be at higher risk for severe COVID-19 [89, 90]. We also detected a marginally significant association between *rs*139227237 and *rs*5743704 [p.(Pro631His)], another missense mutation in *TLR2*, and the incidence of cardiac infarction (P=0.056; **Table S45**). Although this result was not significant after statistical correction, it is notable that the *rs*139227237 C-allele and the *rs*5743704 A-allele were present in individuals who had a cardiac infarction while these alleles were absent in individuals who had not experienced this cardiac event. Additionally, we did not observe any associations between genetic variation at the other immunity genes and comorbidity in this dataset (**Tables S44 and S45**).

We further explored the effect of the coding region on COVID-19 outcomes in an independent dataset. Specifically, we analyzed whole exome sequencing data from a separate cohort of 2,920 SARS-CoV-2-positive individuals from GEN-COVID. These participants were classified by degrees of COVID-19 severity (*i.e.*, not hospitalized, hospitalized with no oxygen therapy, hospitalized with oxygen therapy, hospitalized with CPAP/BiPAP, hospitalized with intubation, and deceased). When we examined these phenotypes as a binary trait (*i.e.*, not-hospitalized versus the more serious outcomes), we detected a significant association at *rs*5743709 in *TLR2* [p.(Ala781Ala)] (P < 0.035; **Table S46**). In addition, when we treated levels of COVID-19 severity as quantitative phenotypes (*i.e.*, as graded levels of COVID-19 severity), we observed a significant association between the *rs*5743709 A-allele and COVID-19 severity (P < 0.008; **Table S47**) using a mixed linear regression model; this linear regression analysis also revealed a small to medium effect size for this variant (**Table S47**). We did not detect any significant associations in the other immunity genes. Lastly, we found that our candidate risk alleles (at *rs*139227237 and *rs*5743709) generally occurred at low frequency in Eurasian populations (between ∼0.05% and ∼1%) but were absent in populations of African descent in the 1000 Genomes Project (**Figure 6**; **Table S2**).

**Figure 6.**
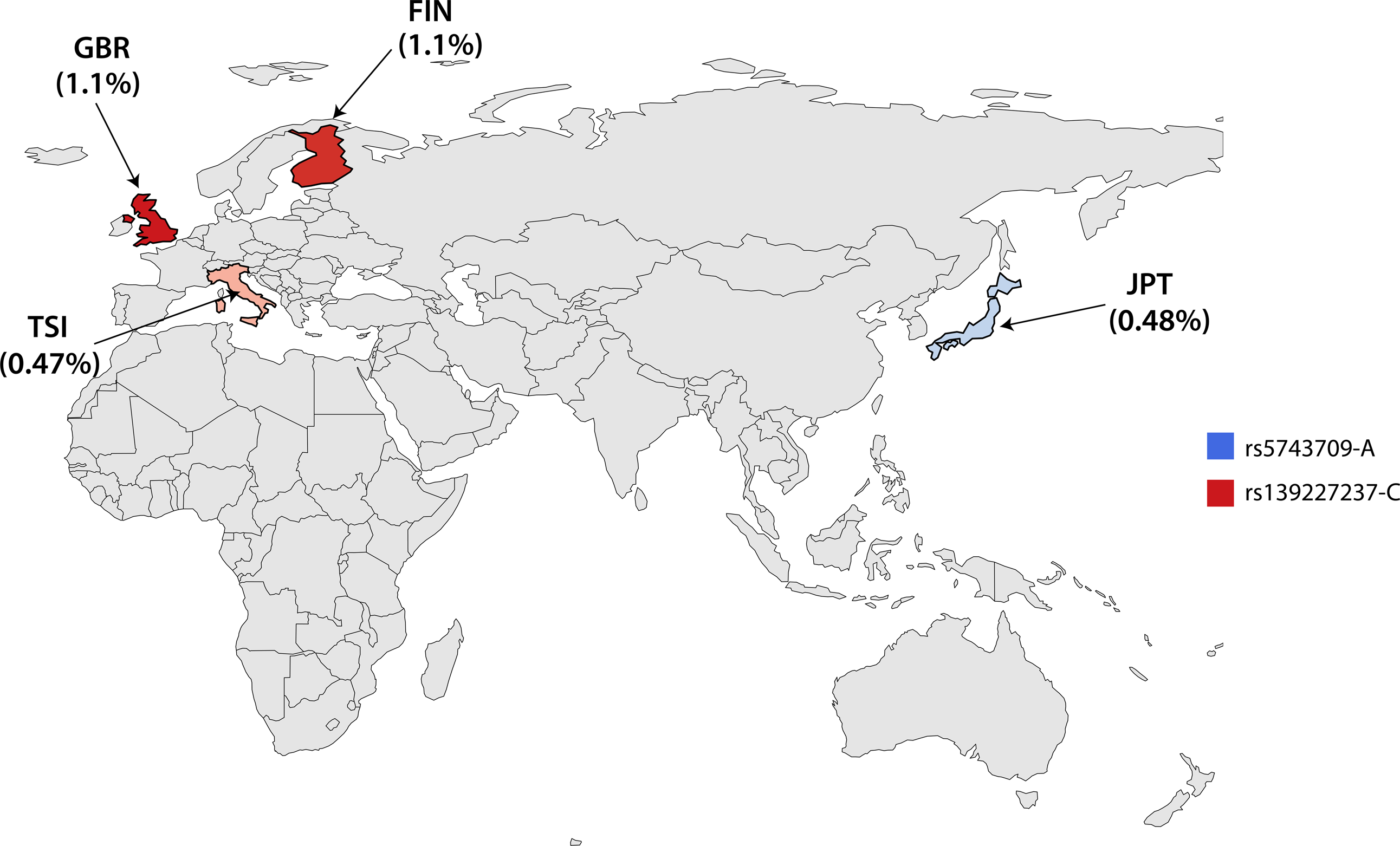
Geographic distribution and the frequency of candidate risk alleles for severe COVID-19. This map displays the geographic distribution of the rs5743709 A-allele and the rs139227237 C-allele in global populations from the 1000 Genomes Project. Particularly, the red shading shows the distribution of the *rs*139227237 C-allele (in the European Great British [GBR], Finnish [FIN], Tuscan Italians [TSI]) while blue shading shows the distribution of the rs5743709 A-allele (in the East Asian Japanese [JPT]). Darker shading reflects a higher allele frequency (∼1.1%) while lighter shading indicates a lower allele frequency (∼0.5%).

## Discussion

### Evidence for positive selection

Our genetic analyses uncovered strong evidence for positive selection at *IL-4*, *TLR2*, *CCL2*, and *SLC11A1* in African and non-African populations. Specifically, we detected significant deviations from neutrality, extended haplotype homozygosity around either derived or ancestral alleles, high levels of population differentiation, and significant *s* estimates at multiple loci at these genes. Taken together, these analyses led to the identification of 72 common SNPs that were inferred to be under selection. Furthermore, several of these SNPs were found on different common haplotypes, suggesting these haplotypes could have risen to high frequency in populations due to multiple-locus soft sweeps [91, 92]. Under this model, no single favored mutation sweeps to fixation [93, 94]; instead beneficial alleles at different nucleotide sites increase in frequency simultaneously undergoing incomplete selective sweeps. In this scenario, multiple alleles (and haplotypes) can co-exist at intermediate frequency, resulting in strongly positive summary statistics in populations, which we observed at *IL-4* (in Africans and non-Africans), *TLR2* (in non-Africans) and *CCL2* (in both Africans and non-Africans).

Among these three genes, some of the most striking genetic patterns arguably occurred in or around *IL-4* which harbored alleles that were inferred targets of selection, including those shared with Neanderthal/Denisovan species—a finding not previously reported in the literature. This result prompts new questions about whether archaic and modern humans displayed similar phenotypic traits due to their shared alleles and whether these alleles were subject to selection in archaic humans. Furthermore, we observed extreme population differentiation primarily at archaic variation at *IL-4* and a pattern of divergent haplotypes. Sakagami et al. (2004), one of the few evolutionary studies of *IL-4*, reported finding divergent haplotypes in their dataset and proposed that this pattern of variation was likely the result of selection in response to different environmental pressures in populations [95]. Overall, our results and those from this previous study highlight the complex nature of adaptive evolution at this gene.

In comparison, the markedly negative *H* values and extensive EHH mainly around derived alleles at *TLR2* in African populations suggest that this gene likely experienced a classic (or hard) selective sweep in Africa. Similar signals of positive selection have been identified at other loci, such as *IFN*-γ on Chromosome 12 [96] and the *LCT* region on Chromosome 2 associated with lactase persistence [77, 97, 98]. Likewise, the pronounced negative *H* values at *SLC11A1* and the long-range haplotype structure around this gene in non-African populations suggest *SLC11A1* underwent a hard selective sweep, leading to the dominance of the H231 haplotype in these groups. Non-Africans also possessed high-frequency selected variants that occurred at low frequency or were absent in populations of African descent, suggesting local (or regional) adaptation outside of Africa. Although African and African-descended populations exhibited significantly negative *H* statistics at *SLC11A1*, they lacked the extensive haplotype structure observed in non-Africans. One possible explanation for this pattern is that classic selective sweeps at this gene in ancestral Africans occurred much earlier in time than those in non-Africans, decreasing the likelihood of observing long-range EHH in African populations. In the end, it is feasible that a skew in the site frequency spectrum indicative of positive selection may be present in modern human populations, but long-range haplotype structure may be absent [99]. In addition, we found haplotypes carrying selected variants that were specific to populations of African descent, likely reflecting local (or regional) adaptation. As the primary interface with the environment, it is not surprising that immunity genes have been subject to local pathogen-driven selection over evolutionary time.

### Functional effects of common adaptive loci

Using the SNP2TFBS tool, we identified four inferred targets of selection that were predicted to alter the binding of multiple transcription factors, potentially affecting gene expression. These transcription factors included EGR2 (rs56174361 and rs56408050 in *IL-4*); SOX10 and ZNF263 (*rs*2857657 in *CCL2*); EGR1 and C/EBPα (*rs*2279015 and rs2279014, respectively, in *SLC11A1*). Previous studies have shown that EGR2 can influence susceptibility to autoimmune eye disorders [100]. Although the function of ZNF263 is not well-understood, this transcription factor has been implicated in basic cellular processes, such as transcriptional repression [101]. SOX10 has been correlated with rare genetic disorders, such as Waardenburg Syndrome [102] and Hirschsprung Disease [103]. Additionally, previous studies have reported that ERG1 regulates cell growth, proliferation, and apoptosis, as well as fibrotic gene expression [104, 105]. Finally, the C/EBPα transcription factor recognizes the CCAAT motif in the promoters of target genes. Collectively, these findings suggest that adaptive loci identified in our study could contribute to gene expression regulation, conferring a fitness advantage.

While computational approaches can be powerful for predicting the impact of nucleotide variation on transcription factor binding [106–112], it is important to recognize their current limitations. In particular, only a small fraction of transcription factors in the genome has been functionally characterized [113]. As a result, not all of the TFBSs, containing our SNPs of interest, have known corresponding transcription factors, limiting our ability to infer function *in silico*. Therefore, future *in vitro* and *in vivo* experiments will be needed to determine the extent to which our loci influence gene expression.

### Relationship between positive selection and risk loci

Interestingly, we found that several common SNPs—previously implicated in respiratory disease risk—were inferred to be under selection. For example, studies have reported that the *rs*13900 C-allele located in the 3′-UTR of *CCL2* was correlated with a decreased level of cytokine production [72, 114], predisposing individuals to infectious disease [115–117]. Our analyses showed that the *rs*13900 T-allele was under selection. Furthermore, *rs*1898830 G-allele in *TLR2* and rs4586 C-allele in *CCL2*, each identified as targets of selection in our study, have been previously implicated in respiratory disease [65, 69–71]. More explicitly, a recent genetic analysis showed that the frequency of the derived *rs*1898830 G-allele was significantly lower in TB patients compared to controls in the Chinese Han population—a finding that was replicated in the Tibetan population [71]. In a separate study, the *rs*4586 T-allele at *CCL2* also was significantly associated with an increased risk for chronic obstructive pulmonary disease, which is characterized by airflow obstruction, chronic inflammation, and a progressive decline in lung function)[65]. Thus, the *rs*13900 T-allele, *rs*1898830 G allele, and the *rs*4586 C allele, all inferred to be under selection, are not associated with increased disease risk and may in fact be protective.

Conversely, we also found that alleles under selection were themselves correlated with increased disease susceptibility. For example, a case-control study of individuals from Japan showed that the derived *rs*2279014 T-allele in the 3’-UTR of *SLC11A1* was associated with protection from *Mycobacterium avium* complex, a predominant contributor to disease burden in the region [118, 119]. However, our results showed selection on the ancestral *rs*2279014 C-allele (rather than the *rs*2279014 T-allele) in East Asian samples, including the Japanese. Furthermore, several studies have reported that the ancestral *rs*2070874 T-allele at *IL-4* was correlated with severe respiratory disease and an exacerbated immune response [57, 66–68]. However, our results indicated selection favoring the ancestral *rs*2070874 T-allele in populations of East Asian and African descent (in contrast, we detected selection on the derived *rs*2070874 C-allele in European and South Asian populations). While it may seem counter-intuitive for alleles contributing to disease susceptibility to be favored in populations, this pattern may be explained by antagonistic pleiotropy—a phenomenon in which a gene or mutation affects multiple traits related to fitness and is antagonistic when a beneficial change in one trait is linked to a disadvantageous change in another within a given environment [120, 121]. Thus, these selected alleles could have been beneficial in one context but costly in another context, particularly as selective pressures change over time. Explicitly, further functional and evolutionary studies are needed to explore this possibility.

### Genotype-phenotype associations

Our analysis of SARS-CoV-2-positive cases from the GEN-COVID Multicenter Study uncovered low-frequency polymorphisms in the *TLR2* coding region that influence COVID-19 and COVID-19-related outcomes. These findings are consistent with other studies that have reported association between host polymorphisms, particularly in genes related to immunodeficiency or inflammasomes, and severe COVID-19 (Elhabyan et al., 2020). For example, mRNA expression levels of Toll-like receptor genes, including *TLR2*, have been associated with disease severity in SARS-CoV-2-infected individuals [19, 39]. Furthermore, rare mutations in the toll-like receptor 7 (TLR-7) gene—which is involved in the innate antiviral immune response—were correlated with severe COVID-19 in patients who were admitted to the ICU in Italy [122]. The authors also found that these mutations suppress the production of interferon, predisposing individuals to severe disease [122]. Similarly, a separate case–control study identified rare missense mutations at *TLR7* in patients with life-threatening COVID-19 [123]. In addition, rare mutations in other immunity genes (e.g., *TLR3*, *IL12RB1*, *TBK1*, and *IFNGR2*) have been argued to play a role in the onset of severe COVID-19 symptoms [122–126]. In contrast, a recent large-scale, multiethnic study of >500,000 individuals found no significant associations between protein-coding variants (ascertained through whole exome sequencing) and COVID-19 severity after applying a stringent Bonferroni correction [127]. However, given this study design, we argue that this analysis would likely not detect any variants with small to medium effect sizes, overlooking novel associations. In contrast, our study focused on alleles that were previously implicated in immune-mediated diseases, allowing us to identify variants with small pleiotropic effects that would otherwise be missed in broader genome-wide analyses. Nonetheless, we recognize that our findings require replication in independent cohorts and/or further functional validation to determine the biological relevance of the variants identified in our study (as well as the ones reported in prior genetic association analyses).

The candidate loci identified in our study have also been implicated in other immune-related diseases in other analyses. For example, data have indicated that the *rs*139227237 [p.(Phe217Ser)] plays a role in abnormal innate immunity signaling and the pathophysiology of myelodysplastic syndromes [128]. Furthermore, the synonymous *rs*5743709 [p.(Ala781Ala)] SNP has been associated with the risk of tuberculosis [129]. Although *rs*5743709 does not alter the amino acid sequence, it is plausible that this SNP could be in strong LD with a functional variant (or exist in a regulatory motif involved in splicing) that directly influences disease risk. Given the overlap in symptoms between severe COVID-19 and other acute illnesses [130, 131], it is possible that the same genetic mutations could affect multiple immune-related diseases (i.e., these alleles could be pleiotropic). Indeed, the pleiotropic effects of variants on disease have been reported in several association studies of complex traits, including disease susceptibility [132–134].

Finally, in contrast to common variants, we found that the low-frequency candidate loci associated with severe COVID-19 were not directly under selection. Given the low frequency of these variants outside of Africa and their absence in our African samples, it is likely these polymorphisms arose more recently following the out-of-Africa migration of modern humans.

## Concluding Remarks

In summary, we uncovered evidence of natural selection and observed the presence of trans-species polymorphisms in modern populations. Furthermore, we identified low-frequency alleles in *TLR2* coding region that are correlated with COVID-19 severity. Although these mutations likely explain a small fraction of COVID-19 variability, a more detailed study of this genetic variation—and other low-frequency variants of small to medium effect—could lead to new insights into the pathophysiology of severe COVID-19. Given the persistence of COVID-19, there is a growing need to identify additional predictive biomarkers of risk for severe disease. In addition to acute disease, long COVID, which is characterized by lingering symptoms affecting multiple organ systems, remains a poorly understood and understudied consequence of SARS-CoV-2 infection. Expanding genetic studies to identify variants associated with post-viral syndromes, such as long COVID, may reveal important pathways underlying chronic immune dysregulation and recovery.

As humans continue to transform their environments, emerging respiratory diseases and their global spread will be one of the grand challenges that countries around the world will face over the coming decades. To minimize the impact of these respiratory illnesses, novel statistical methods and a concerted effort to include diverse populations in genomic studies will be needed. Such approaches will be essential for identifying additional risk alleles that influence disease susceptibility and severity in different populations. Ultimately, the findings from these studies will deepen our understanding of the genetic architecture underlying differential infectious disease risk and help guide the development of effective therapeutic interventions.

## Methods

### Population samples

We analyzed nucleotide and haplotype variation in four immune-related genes: *IL*-*4* (8,698 base pairs [bps]) on chromosome 5, *TLR2* (22,623 bps) on chromosome 4, *CCL2* (2,293 bps) on chromosome 17, and *SLC11A1* (14,926 bps) on chromosome 2 in 2,039 unrelated individuals from the 1000 Genomes Project (Phase 3). These individuals originated from Africa, South Asia, Europe and East Asia [135]. In this dataset, Africa consisted of 99 Esan (ESN, Nigeria), 113 Mandinka (GWD, Gambia), 54 Mende (MSL, Sierra Leone), 107 Yoruba (YRI, Nigeria), 99 Luhya (LWK, Kenya), 60 African Americans (ASW, United States), and 96 Barbadians (ACB, Barbados). Throughout this paper, the term “African populations” refers to indigenous African populations only while the term “African and African-descended populations” refers to indigenous Africans and African Americans/Afro-Caribbean populations, respectively. In addition, we periodically refer to these populations collectively as populations of African descent. South Asia included 84 Bengali (BEB, Bangladesh), 103 Gurjarati (GIH, Gujarati Indian recruited in Houston, Texas), 99 Indian Telugu (ITU, Indian Telugu recruited in the United Kingdom), 93 Punjabi (PJL, Pakistan), and 99 Tamil (STU, Sri Lanka). Europe consisted of 90 Great British (GBR, Great Britain), 99 Finnish (FIN, Finland), 107 Iberian (IBS, Spain), and 104 Toscani Italians (TSI, Italy). East Asia encompassed 93 Dai (CDX, China), 103 Han (CHB, Han Chinese in Beijing, China), 105 Han (CHS, Han Chinese in South China), 104 Japanese (JPT, Japan), and 99 Kinh (KHV, Vietnam). Individual chromosomes for the Altai Neanderthal genome were downloaded from http://cdna.eva.mpg.de/neandertal/altai/AltaiNeandertal/VCF/. Individual chromosomes for the Denisovan genome were downloaded from http://cdna.eva.mpg.de/denisova/VCF/human/.

To investigate the genetic architecture of COVID-19 severity, we examined genetic and clinical data from GEN-COVID, a multicenter network of more than 40 hospitals and affiliated partners that collected high-quality samples and data across Italy for COVID-19 research under the direction of Dr. Alesandra Renieri. More specifically, we analyzed variants genotyped with the Illumina Global Screening Array-24 v3.0 + Multi-Disease BeadChip together with traits related to COVID-19 severity in a cohort of 1,141 SARS-CoV-2-infected participants. In addition, we analyzed whole-exome sequencing data and graded levels of COVID-19 severity in a different cohort of 2,920 SARS-CoV-2-positive individuals from GEN-COVID. The whole exome data were generated using the Illumina NovaSeq6000 System (Illumina, San Diego, CA, USA) with ≥ 20 x coverage. Variant calling was also performed using the GATK best practice guidelines. Notably, there is no overlap of SARS-CoV-2-positive individuals between the genotyping and whole exome sequencing datasets.

### Nucleotide variation

We extracted variant calls (from vcf files; build GRCh37 human assembly) for the *IL*-*4, TLR2, CCL2,* and *SLC11A1* genes in the 1000 Genomes using vcftools [136]. The genomic coordinates (*i.e.*, the start and end positions) for these genes were obtained from the NCBI database. The minor allele frequency (MAF) was defined as the second most frequent allele at a given site in pooled populations (*i.e.*, 2N=4078). SNPs were also broadly classified as common or rare based on the MAF at a given site. More explicitly, SNPs with an MAF ≥ 5% were classified as common, while SNPs with an MAF < 5% were classified as rare.

### Tests of neutrality

To determine if *IL*-*4, TLR2, CCL2,* and *SLC11A1* evolved neutrally, we calculated two summary statistics, Fay and Wu’s *H* (*H*)[137] and Tajima’s *D* (*D*_T_)[138], for each population using DnaSP [75]. These statistical tests analyze different aspects of the site frequency spectrum [139], providing information about the evolutionary forces that have influenced patterns of polymorphic variation in populations. We also simulated theoretical *H* and *D*_T_ values within a coalescent framework under different demographic models [140–145] using the msprime software [146, 147]. For African populations, we simulated a 2–, 4–, 6–, 8–, and 10– fold increase in population size (from an initial population size of 7,300 individuals) starting at 148,000 years ago (ya) until the present (2,000 replicates per simulation).

For non-Africans, we simulated demographic models involving a population bottleneck (consisting of 1,861 individuals) 51,000 years ago, which corresponds to the approximate time anatomically modern humans migrated out of Africa [143, 148, 149], followed by a 20–, 40–, 60–, 80–, and 100– fold increase in population size starting at 23,000 years ago until the present (2,000 replicates per simulation)[140, 141, 143–145, 150]. For all simulations, we used a mutation rate of 2.5 x 10^-8^, recombination rate of 1 x 10^-8^, and a generation time of 28 years [143–145]. We also incorporated bidirectional migration between Africans and non-Africans at ∼30,000 ya and ∼20,000 ya using the number of migrants given in Gravel et al. [143]. Finally, we assessed the statistical significance of observed *H* and *D*_T_ values by comparing them to a distribution of theoretical values. In cases where observed values were either negative or positive in populations, we tested for significance in one direction (*i.e.*, a one-tailed test, *P* < 0.05). In cases where observed values were a mix of positive and negative numbers in populations, we tested for significant difference in both directions (*i.e.*, a two-tailed test, *P* < 0. 05).

### Long-range haplotype homozygosity

We characterized the length of haplotype homozygosity on Chromosomes 2 (*SLC11A1*), 4 (*TLR2*), 5 (*IL-4*), and 17 (*CCL2*) for each population using the *i*HS statistic calculated with the selscan and Polaris packages [76, 142, 151]. To identify outlier *i*HS values, the unstandardized scores for >600,000 SNPs across the ∼240 Mb region of Chromosome 2, >520,000 SNPs across the ∼190 Mb region of Chromosome 4, >470,000 SNPs across the ∼180 Mb region of Chromosome 5, and >180,000 SNPs across the ∼80 Mb region of Chromosome 17 in each population were normalized using the norm program implemented in selscan [76]. Standardized | *i*HS | > 2 represents the top 5% of | iHS | scores. We also calculated *n*S_L_, another haplotype-based statistic, to estimate the length of haplotype homozygosity in populations using selscan [152]. Unlike the *i*HS, this statistic measures haplotype lengths based on the number of segregating sites in a sample and does not depend on the recombination rate [152], making it robust to recombination rate variation. Furthermore, *nS*_L_ can detect selective sweeps from standing variation and incomplete selective sweeps [152]. The resulting *nS*_L_ statistics were normalized using the norm software [76]. By convention, selscan reports positive statistics when the derived allele (coded as “1”) is under selection and negative statistics when the ancestral allele (coded as “0”) is favored [76, 153–155], which is opposite to the orientation described in Voight *et al.* [142].

Finally, we quantified the decay of identity of haplotypes with distance by calculating the extended haplotype homozygosity (EHH) statistic with selscan and Polaris [76, 78, 151] using loci with extreme | iHS | and/or | *nS*_L_ | scores identified above as core SNPs. If we observed outlier standardized | iHS | and/or | *nS*_L_ | statistics along with long-range EHH for a given SNP, we classified that SNP as a target of selection.

### Selection coefficient estimates

The selection coefficient (*s*) quantifies the fitness effects of mutations that arise in populations, providing insight into the impact of selection at the genetic level. We estimated *s* for alleles with an MAF ≥ 5% in our immunity genes using the CLUES2 algorithm implemented in the CLUES2 Companion package [79, 81]. CLUES2 incorporates ancestral recombination graphs (ARGs)—which capture the history of genomic regions in a sample of sequences—along with importance sampling of ARGs to estimate *s* in different time periods [79]. Thus, CLUES2 utilizes the full set of allele histories in a sample of sequences to estimate *s* [79]. We calculated *s* at different time depths (i.e., 1,000, 500, and 200 generations ago, which correspond to 28,000, 14,000, and 5,600 ya assuming a generation time of 28 years) in each gene using CLUES2 Companion [81]. Because selection intensity can vary over time [79], we aimed to capture time points at which selection coefficients might change. SNP loci with significant *s* estimates (i.e., −log(p) > 1.3 or *P* < 0.05) at any time depth were classified as targets of positive selection, complementing the | iHS |, | *nS*_L_ |, and EHH results.

### Population differentiation

To measure the degree of genetic divergence among populations, we calculated among-population *F*_ST_ at individual polymorphic sites across each gene using the Weir and Cockerham method in vcftools [136, 156]. The observed *F*_ST_ estimates for SNPs in the *IL-4*, *TLR2*, *CCL2*, and *SLC11A1* genes were then compared against an empirical distribution of *F*_ST_ values derived from 8.1 million genome-wide SNPs that were randomly chosen to identify outlier values (≥ 95th percentile of the distribution). This *F*_ST_ empirical distribution was generated using the same 21 populations that were used to calculate the observed values. Based on this approach, we determined that *F*_ST_ > 0.286 represented the top 95^th^ percentile of empirical values.

### Haplotype inferences and relationships

Fully phased haplotypes from the 1000 Genomes Project were extracted for 2,039 individuals using vcftools [136]. A custom script was applied to the already phased sequence data to identify unique haplotypes along with the count of each haplotype in the dataset. Next, the genealogical relationships among haplotypes were reconstructed using the median-joining method implemented in the Network v. 5.0 software [82]. For each gene, the resulting visual tree for each gene was the one with the least number of changes among all possible trees [157].

### *In silico* functional analysis

We applied the SNP2TFBS tool to polymorphisms that were inferred to be under selection [87]. This tool assesses whether or not SNPs of interest map to transcription factor binding sites (TFBSs) and outputs a list of SNPs that overlap with TFBSs along with the transcription factors that interact with them [87].

### Genetic association analyses

We examined genotypic and clinical data from laboratory-confirmed SARS-CoV-2-postitive individuals enrolled in the GEN-COVID Multicenter Study in Italy. The GEN-COVID cohorts consisted of SARS-CoV-2-infected individuals from whom DNA and detailed clinical information were collected through a consortium of more than 40 Italian Hospitals, 16 Continuity Assistance Special Units, and eight Departments of Preventive Medicine for host genetic analysis.

In our first GEN-COVID cohort of 1,141 SARS-CoV-2-positive individuals, we analyzed comorbidities (specifically, cancer, chronic kidney, chronic pulmonary, transplant, heart failure, diabetes, immunocompromised, asthma, hypertension, cardiac infarction, stroke, and ICU admission status) known to increase risk for severe COVID-19 along with genotypes from our four genes. This dataset consisted of 648 males and 493 females ranging in age from 18 to 99 years. Prior to testing for any genetic associations, we generated a genetic relationship matrix (GRM) between pairs of individuals using genome-wide SNPs in the 1,141 SARS-CoV-2-positive individuals to account for population structure and familial relatedness with the GTCA software [158]. We then used a linear mixed model approach implemented in GCTA [158] to test for associations between genetic variation and traits related to severe COVID-19 in our cohort (sex and age were included in this analysis as covariates). Statistical significance was determined after applying: 1) the saddle point approximation (SPA) method to correct for unbalanced samples sizes in comparative groups; and 2) the Benjamini-Hochberg method to correct for multiple testing, mitigating p-value inflation. Additionally, we analyzed whole exome sequencing data in a second GEN-COVID cohort comprised of 2,920 SARS-CoV-2-infected individuals (1,730 males and 1,189 females ranging in age from a few months to 99 years). These 2,920 SARS-CoV-2-positive cases were also classified by degrees of COVID-19 severity; specifically, score 0=not hospitalized (N=499), 1=hospitalized with no oxygen therapy (N=406), 2=hospitalized with oxygen therapy (N=946), 3=hospitalized with CPAP/BiPAP intervention (N=668), 4=hospitalized with intubation (N=207), and 5=deceased (N=194). Severe COVID-19 in the current study is defined as the stage of illness requiring hospitalization or resulting in death. For this analysis, we generated a new GRM using the whole exome sequencing data from our 2,920 SARS-CoV-2-positive cases. We then employed linear mixed model approaches implemented in GCTA [158] to examine the effects of exonic variants on COVID-19 severity. More specifically, we analyzed phenotypes as binary and quantitative traits using logistic and linear methods, respectively. In both analyses, the GRM was incorporated as a random variable to account for population structure and familial relatedness; sex and age were included as covariates. Statistical significance was determined after applying SPA and Benjamini-Hochberg corrections. It is important to note that there was no overlap of individuals between the first and second cohorts of SARS-CoV-2-infected individuals.

## Acknowledgements

We thank the Center for Computational Biology and Bioinformatics (CCBB) at Howard University and the Center for Advanced Research Computing at the University of Southern California for providing computational resources essential to this study. We are also grateful to Professor Alessandra Renieri at the University of Siena (Italy) for providing the genotype–phenotype data used in our analyses.

## Author contributions

All authors contributed to the study conception and design. Material preparation and analyses were performed by Christopher N. Cross, Alessandro Lisi, Faith C. Simmonds, and Michael C. Campbell. The first draft of the manuscript was written by Christopher N. Cross and all authors commented on previous versions of the manuscript. All authors read and approved the final manuscript.

## Data availability

The genetic diversity data underlying this article are available in the 1000 Genomes Consortium Project at https://ftp.1000genomes.ebi.ac.uk/vol1/ftp/release/20130502/. The clinical datasets analyzed for the current study are available from GEN-COVID upon reasonable request.

## Abbreviations

ACB: African Caribbeans in Barbados
ACE2: Angiotensin-Converting Enzyme 2
ANPEP: Alanyl Aminopeptidase
APC: Antigen-Presenting Cell
ARG: Ancestral Recombination Graph
ASW: African Ancestry in Southwest US
BEB: Bengali in Bangladesh
bp: Base Pair
CCL2: C-C Motif Chemokine Ligand 2
CCL3: C-C Motif Chemokine Ligand 3
CCBB: Center for Computational Biology and Bioinformatics
CDX: Chinese Dai in Xishuangbanna, China
CHB: Han Chinese in Beijing, China
CHS: Southern Han Chinese
CLUES2: Composite Likelihood for Estimating Selection, version 2
COVID-19: Coronavirus Disease 2019
CPAP: Continuous Positive Airway Pressure
CTSB: Cathepsin B
CTSL: Cathepsin L
CXCL10: C-X-C Motif Chemokine Ligand 10
DPP4: Dipeptidyl Peptidase-4
DNA: Deoxyribonucleic Acid
D_T_: Tajima’s D Statistic
EHH: Extended Haplotype Homozygosity
ER: Endoplasmic Reticulum
ESN: Esan in Nigeria
FIN: Finnish in Finland
F_ST_: Fixation Index
FURIN: Furin Protease
GBR: British in England and Scotland
GEN-COVID: Genetic and Clinical Data Collection for COVID-19
GIH: Gujarati Indian in Houston, Texas
GRCh37: Genome Reference Consortium Human Build 37
GRM: Genetic Relationship Matrix
GWD: Gambian in Western Division, The Gambia
H: Fay and Wu’s H Statistic
HBV: Hepatitis B Virus
HCV: Hepatitis C Virus
HIV: Human Immunodeficiency Virus
HLA: Human Leukocyte Antigen
IBD: Inflammatory Bowel Disease
IBS: Iberian population in Spain
ICU: Intensive Care Unit
IFN-α: Interferon-alpha
IFN-γ: Interferon-gamma
iHS: Integrated Haplotype Score
IL: Interleukin
IL-4: Interleukin-4
IL-6: Interleukin-6
IL-10: Interleukin-10
ITU: Indian Telugu in the UK
JPT: Japanese in Tokyo, Japan
KHV: Kinh in Ho Chi Minh City, Vietnam
LD: Linkage Disequilibrium
LWK: Luhya in Webuye, Kenya
MAF: Minor Allele Frequency
MSL: Mende in Sierra Leone
mRNA: Messenger RNA
nS_L_: Number of Segregating Sites by Length (haplotype-based test)
ORF: Open Reading Frame
PCA: Principal Component Analysis
PJL: Punjabi in Lahore, Pakistan
RNA: Ribonucleic Acid
SARS-CoV-2: Severe Acute Respiratory Syndrome Coronavirus 2
SLC11A1: Solute Carrier Family 11 Member 1
SNP: Single Nucleotide Polymorphism
SPA: Saddle Point Approximation
STU: Sri Lankan Tamil in the UK
TB: Tuberculosis
TF: Transcription Factor
TFBS: Transcription Factor Binding Site
TLR: Toll-like Receptor
TLR2: Toll-like Receptor 2
TMPRSS2: Transmembrane Protease, Serine 2
TMPRSS11D: Transmembrane Protease, Serine 11D
TSI: Toscani in Italy
UTR: Untranslated Region
VCF: Variant Call Format
YRI: Yoruba in Ibadan, Nigeria

