## Supplementary material for "The evolutionary landscape of host immunity genes involved in respiratory and other immune-related diseases, and their association with severe COVID-19 outcomes": Figure S3

**Figure S3: Extended Haplotype Homozygosity (EHH) around *IL-4* on Chromosome 5 in global populations.** EHH plots around allelic variation around *IL-4* show the decay of identity of haplotypes on chromosomes. The blue shading shows the decay of homozygosity of chromosomes carrying the ancestral allele at the core, while the red shading indicates the decay of homozygosity on chromosomes with the derived allele at the core site in different populations from the 1000 Genomes Project. The distance from the core SNP (at zero) is displayed on the x-axis; the negative numbers indicate distance upstream from the core SNP, while positive values indicate distance downstream from the core SNP on the forward strand. The EHH probabilities are shown on the y-axis.

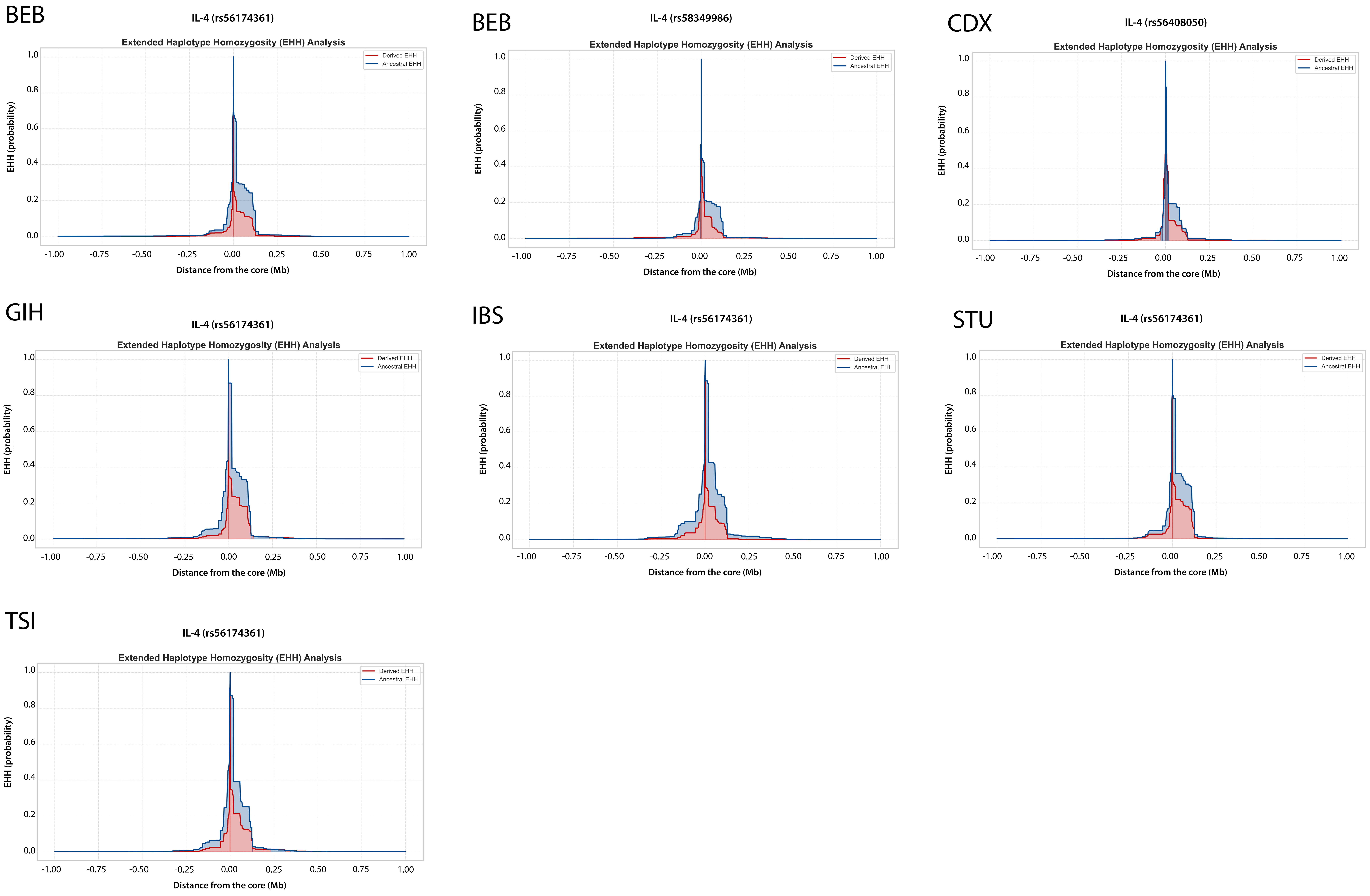
BEB
IL-4 (rs56174361)
BEB
IL-4 (rs58349986)
CDX
IL-4 (rs56408050)
GIH
IL-4 (rs56174361)
IBS
IL-4 (rs56174361)
STU
IL-4 (rs56174361)
TSI
IL-4 (rs56174361)
Extended Haplotype Homozygosity (EHH) Analysis
Derived EHH
Ancestral EHH
EHH (probability)
Distance from the core (Mb)
1.0
0.8
0.6
0.4
0.2
0.0
-1.00
-0.75
-0.50
-0.25
0.00
0.25
0.50
0.75
1.00

**Figure S3**
