## Supplementary material for "The evolutionary landscape of host immunity genes involved in respiratory and other immune-related diseases, and their association with severe COVID-19 outcomes": Figure S6

**Figure S6: Extended Haplotype Homozygosity (EHH) around *TLR2* on Chromosome 4 in global populations.** EHH plots around allelic variation around *TLR2* show the decay of identity of haplotypes on chromosomes. The blue shading shows the decay of homozygosity of chromosomes carrying the ancestral allele at the core, while the red shading indicates the decay of homozygosity on chromosomes with the derived allele at the core site in different populations from the 1000 Genomes Project. The distance from the core SNP (at zero) is displayed on the x-axis; the negative numbers indicate distance upstream from the core SNP, while positive values indicate distance downstream from the core SNP on the forward strand. The EHH probabilities are shown on the y-axis.

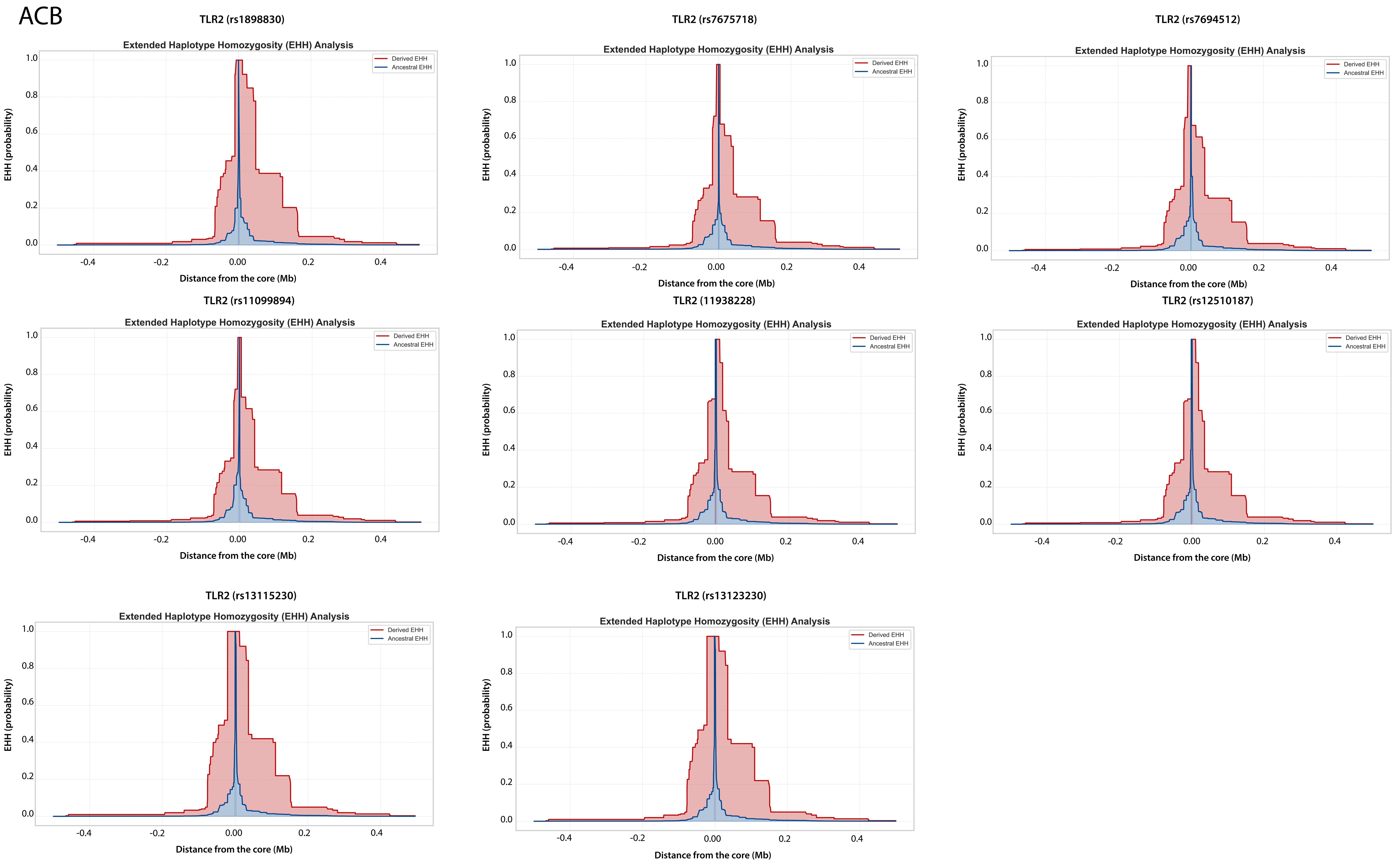
ACB
TLR2 (rs1898830)
TLR2 (rs7675718)
TLR2 (rs7694512)
TLR2 (rs11099894)
TLR2 (11938228)
TLR2 (rs12510187)
TLR2 (rs13115230)
TLR2 (rs13123230)
Extended Haplotype Homozygosity (EHH) Analysis
Derived EHH
Ancestral EHH
EHH (probability)
Distance from the core (Mb)
1.0
0.8
0.6
0.4
0.2
0.0
-0.4
-0.2
0.00
0.2
0.4

**Figure S6**

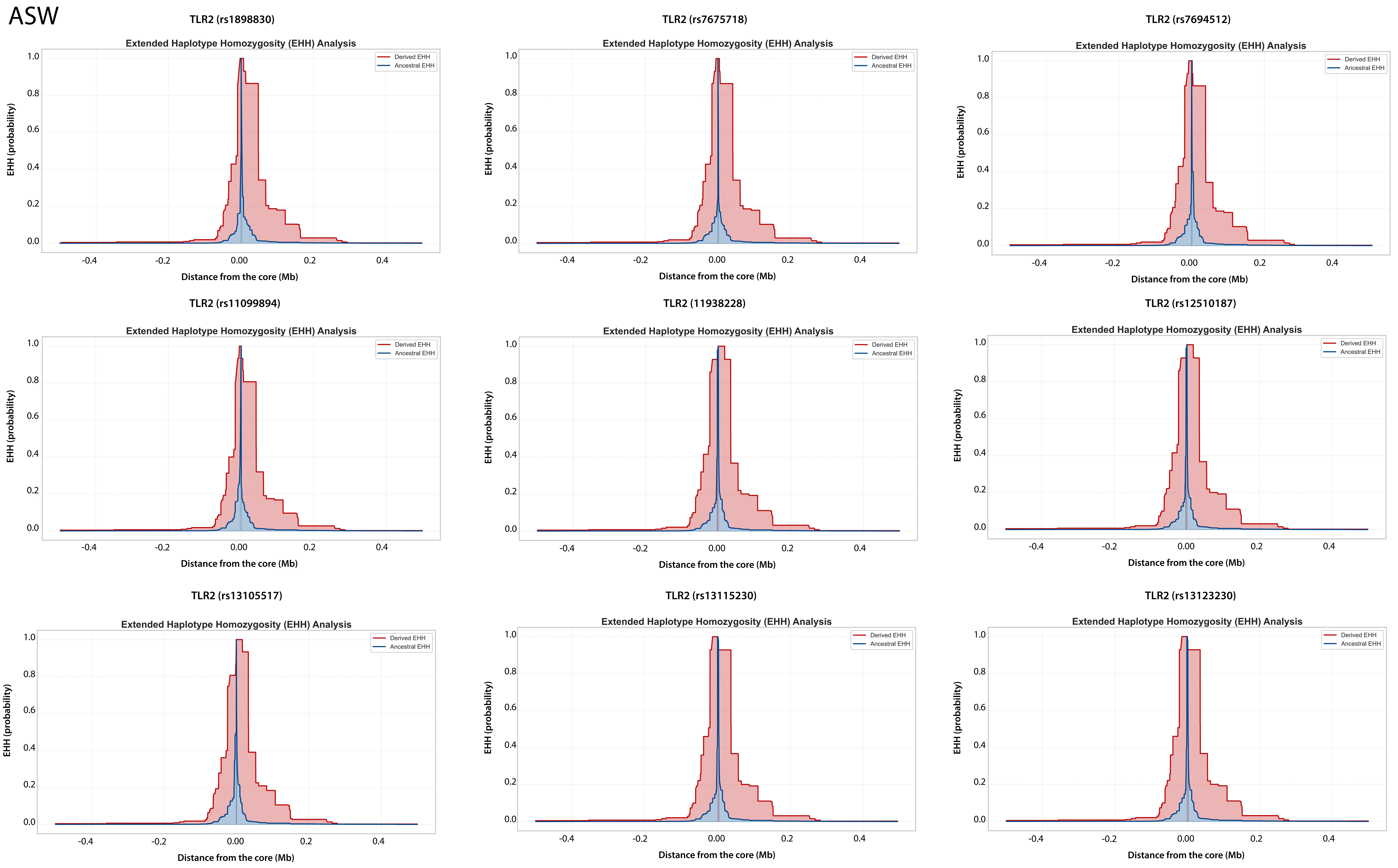
ASW
TLR2 (rs1898830)
TLR2 (rs7675718)
TLR2 (rs7694512)
TLR2 (rs11099894)
TLR2 (11938228)
TLR2 (rs12510187)
TLR2 (rs13105517)
TLR2 (rs13115230)
TLR2 (rs13123230)
Extended Haplotype Homozygosity (EHH) Analysis
Derived EHH
Ancestral EHH
EHH (probability)
Distance from the core (Mb)
1.0
0.8
0.6
0.4
0.2
0.0
-0.4
-0.2
0.00
0.2
0.4

**Figure S6**

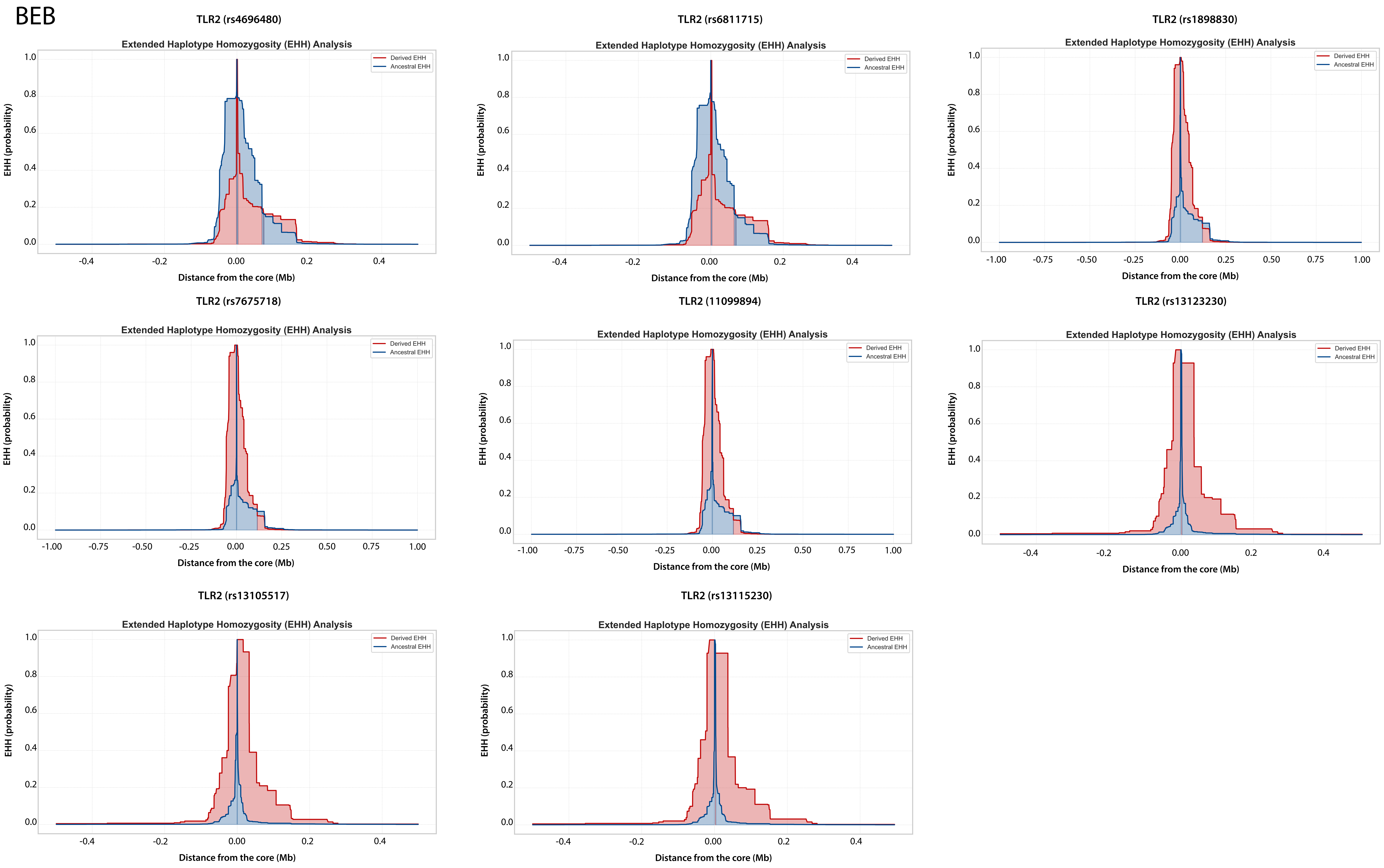
BEB
TLR2 (rs4696480)
TLR2 (rs6811715)
TLR2 (rs1898830)
TLR2 (rs7675718)
TLR2 (11099894)
TLR2 (rs13123230)
TLR2 (rs13105517)
TLR2 (rs13115230)
Extended Haplotype Homozygosity (EHH) Analysis
Derived EHH
Ancestral EHH
EHH (probability)
Distance from the core (Mb)

### Figure S6

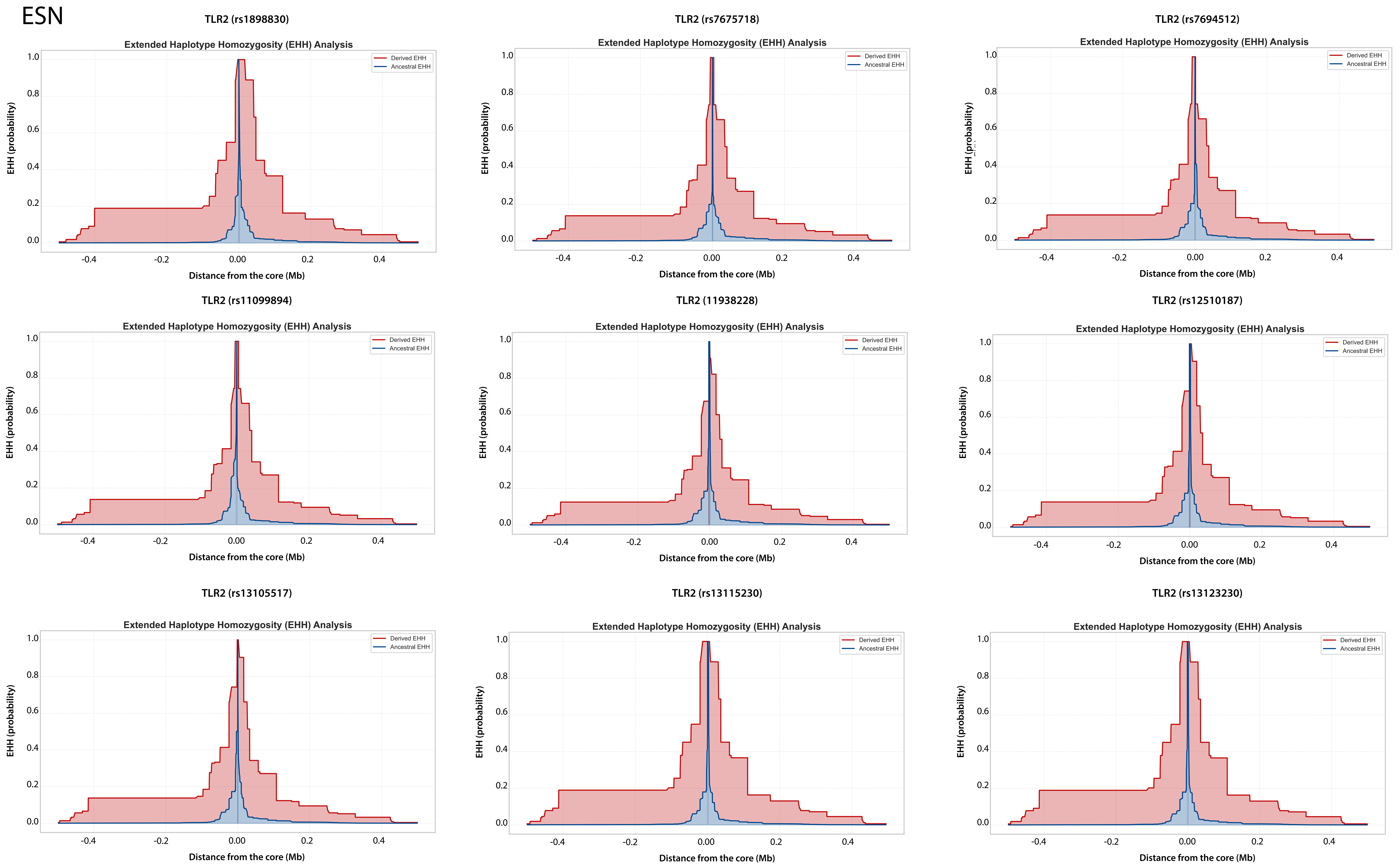
ESN
TLR2 (rs1898830)
TLR2 (rs7675718)
TLR2 (rs7694512)
TLR2 (rs11099894)
TLR2 (11938228)
TLR2 (rs12510187)
TLR2 (rs13105517)
TLR2 (rs13115230)
TLR2 (rs13123230)
Extended Haplotype Homozygosity (EHH) Analysis
Derived EHH
Ancestral EHH
EHH (probability)
Distance from the core (Mb)
1.0
0.8
0.6
0.4
0.2
0.0
-0.4
-0.2
0.00
0.2
0.4

**Figure S6**

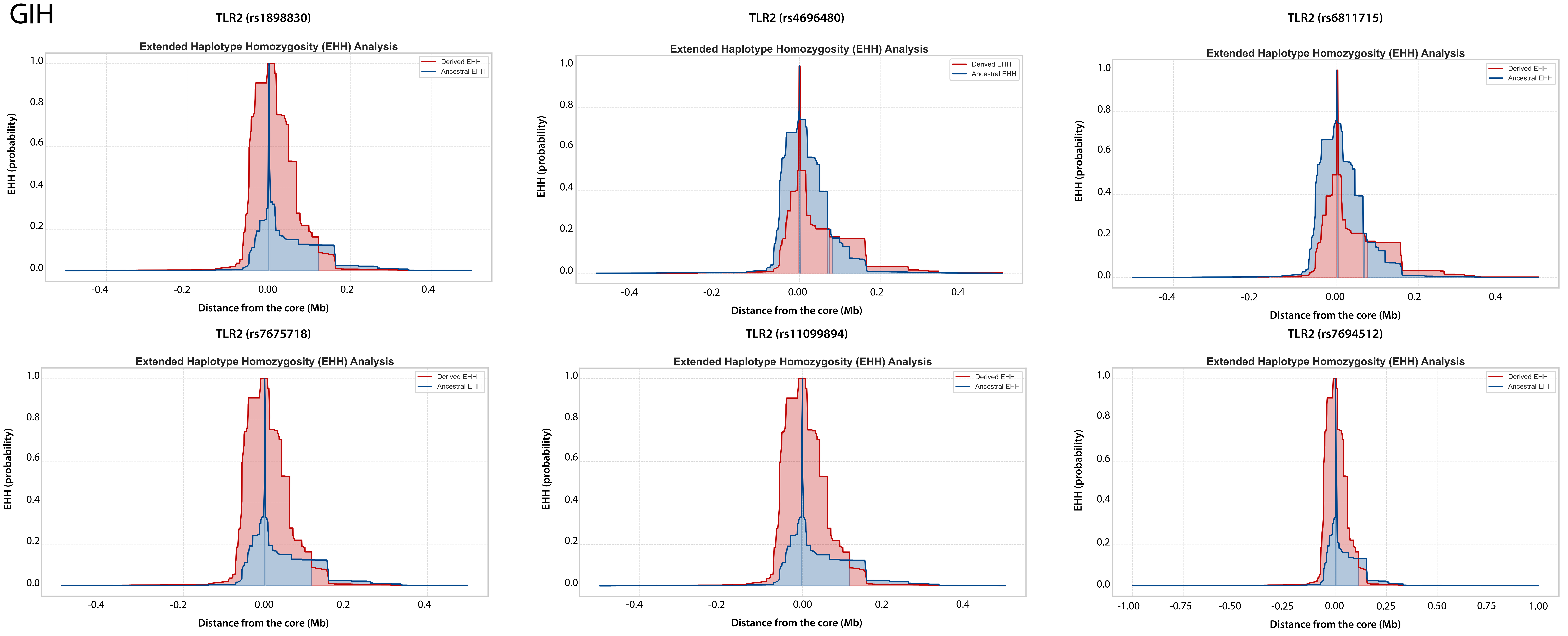
GIH
TLR2 (rs1898830)
TLR2 (rs4696480)
TLR2 (rs6811715)
TLR2 (rs7675718)
TLR2 (rs11099894)
TLR2 (rs7694512)
Extended Haplotype Homozygosity (EHH) Analysis
Derived EHH
Ancestral EHH
EHH (probability)
Distance from the core (Mb)

**Figure S6**

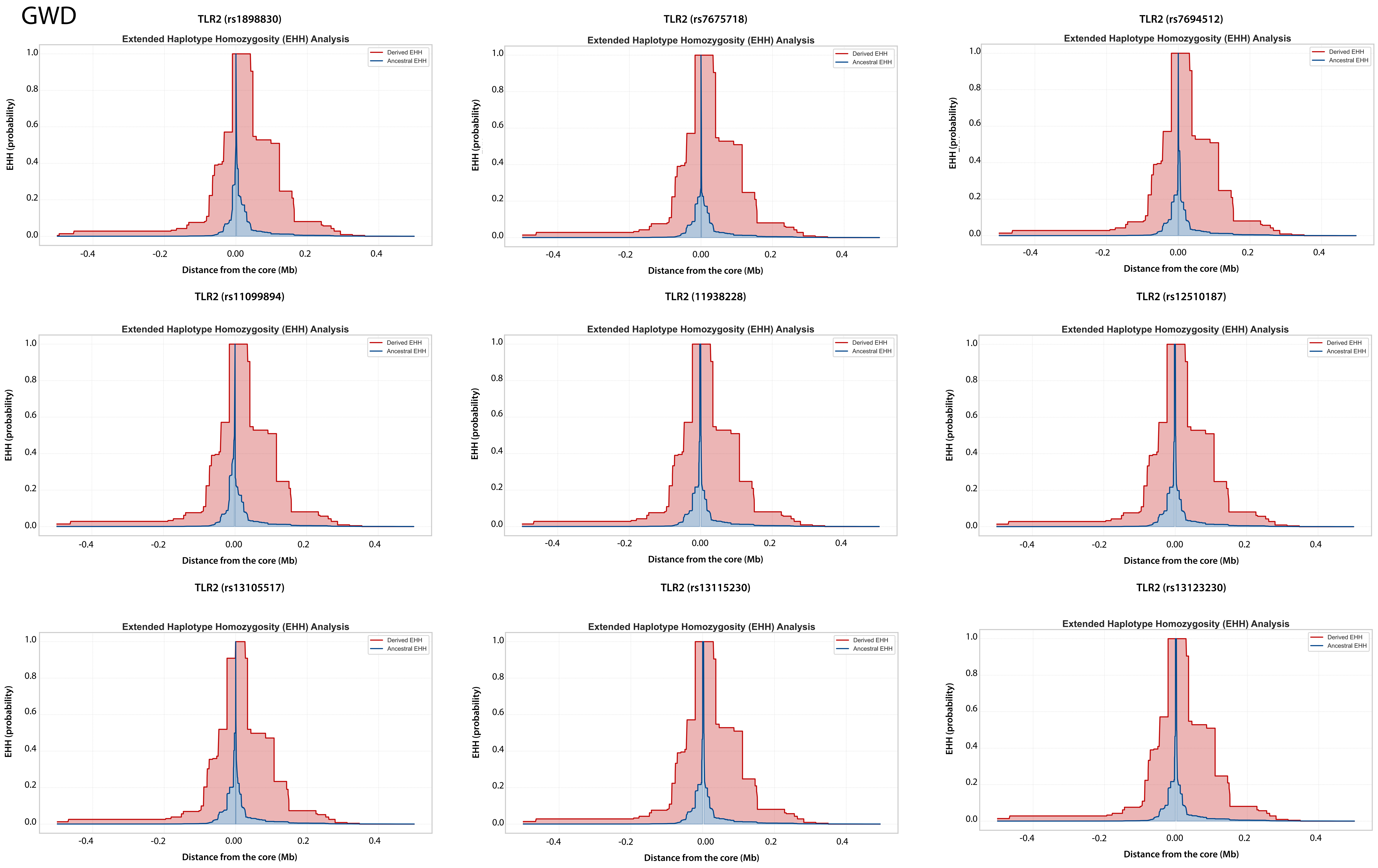
GWD
TLR2 (rs1898830)
TLR2 (rs7675718)
TLR2 (rs7694512)
TLR2 (rs11099894)
TLR2 (11938228)
TLR2 (rs12510187)
TLR2 (rs13105517)
TLR2 (rs13115230)
TLR2 (rs13123230)
Extended Haplotype Homozygosity (EHH) Analysis
Derived EHH
Ancestral EHH
EHH (probability)
Distance from the core (Mb)
1.0
0.8
0.6
0.4
0.2
0.0
-0.4
-0.2
0.00
0.2
0.4

**Figure S6**

ITU

TLR2 (rs4696480)

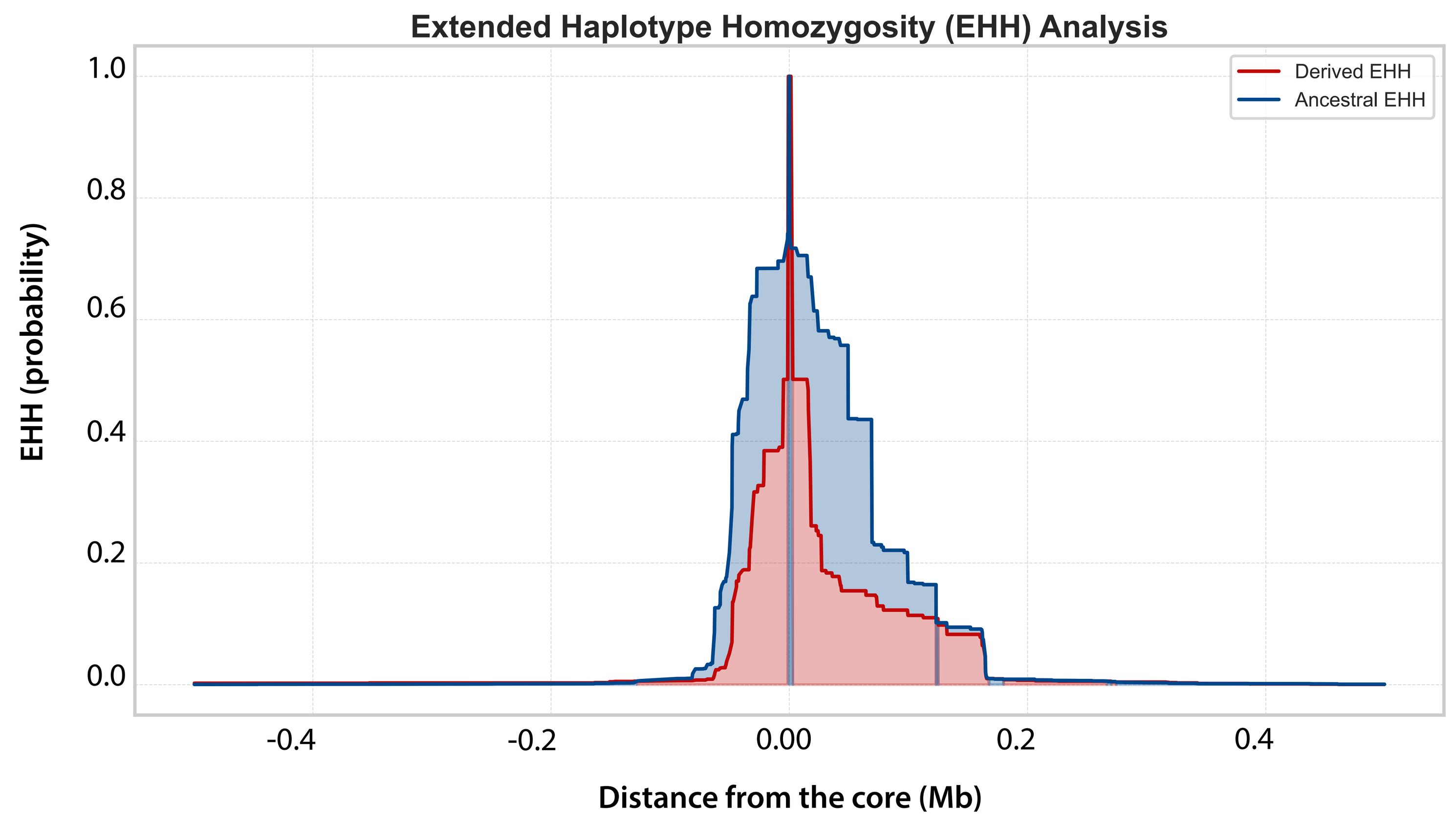


TLR2 (rs6811715)

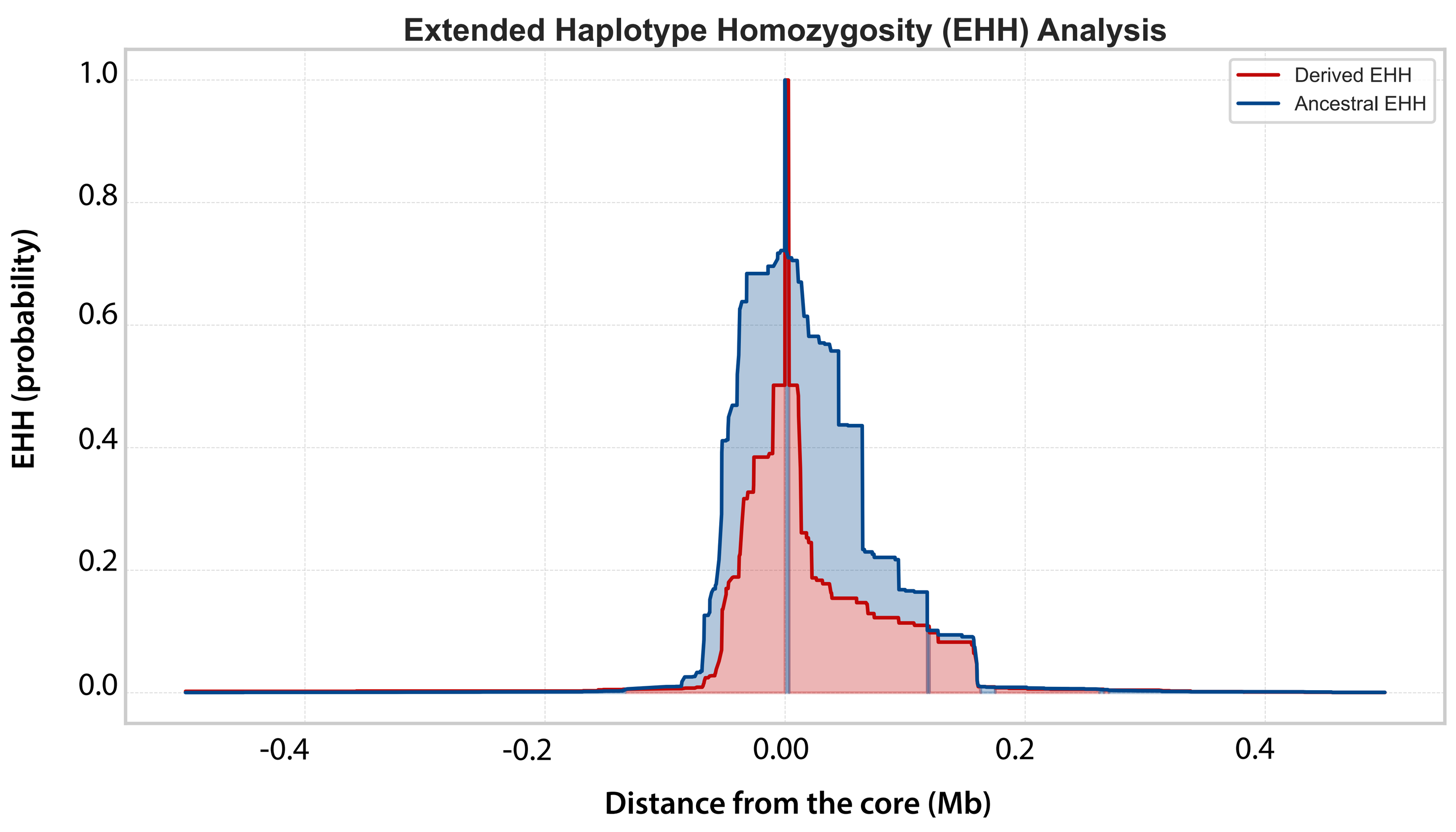


TLR2 (rs1898830)

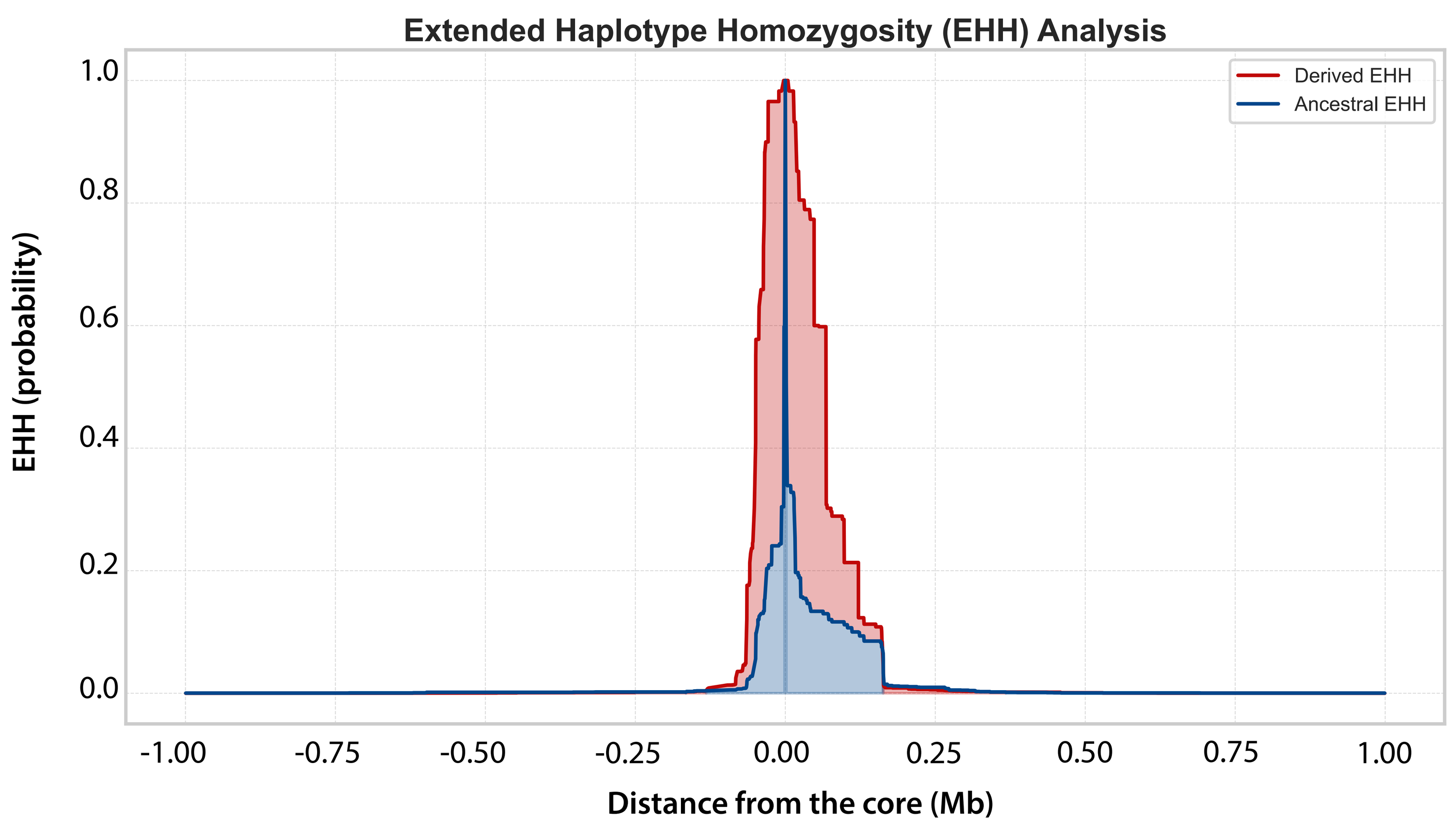


TLR2 (rs7675718)

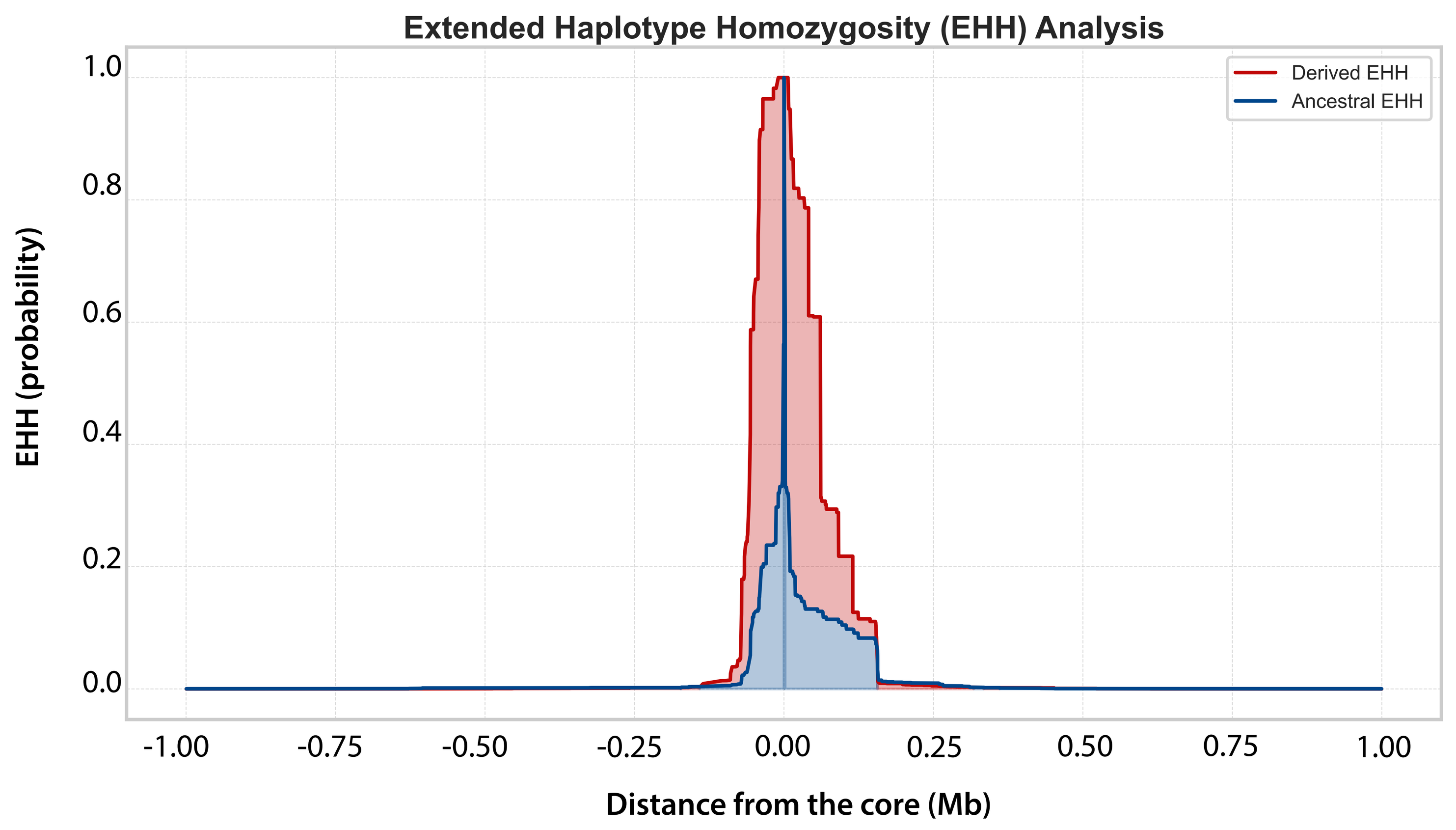


TLR2 (rs7694512)

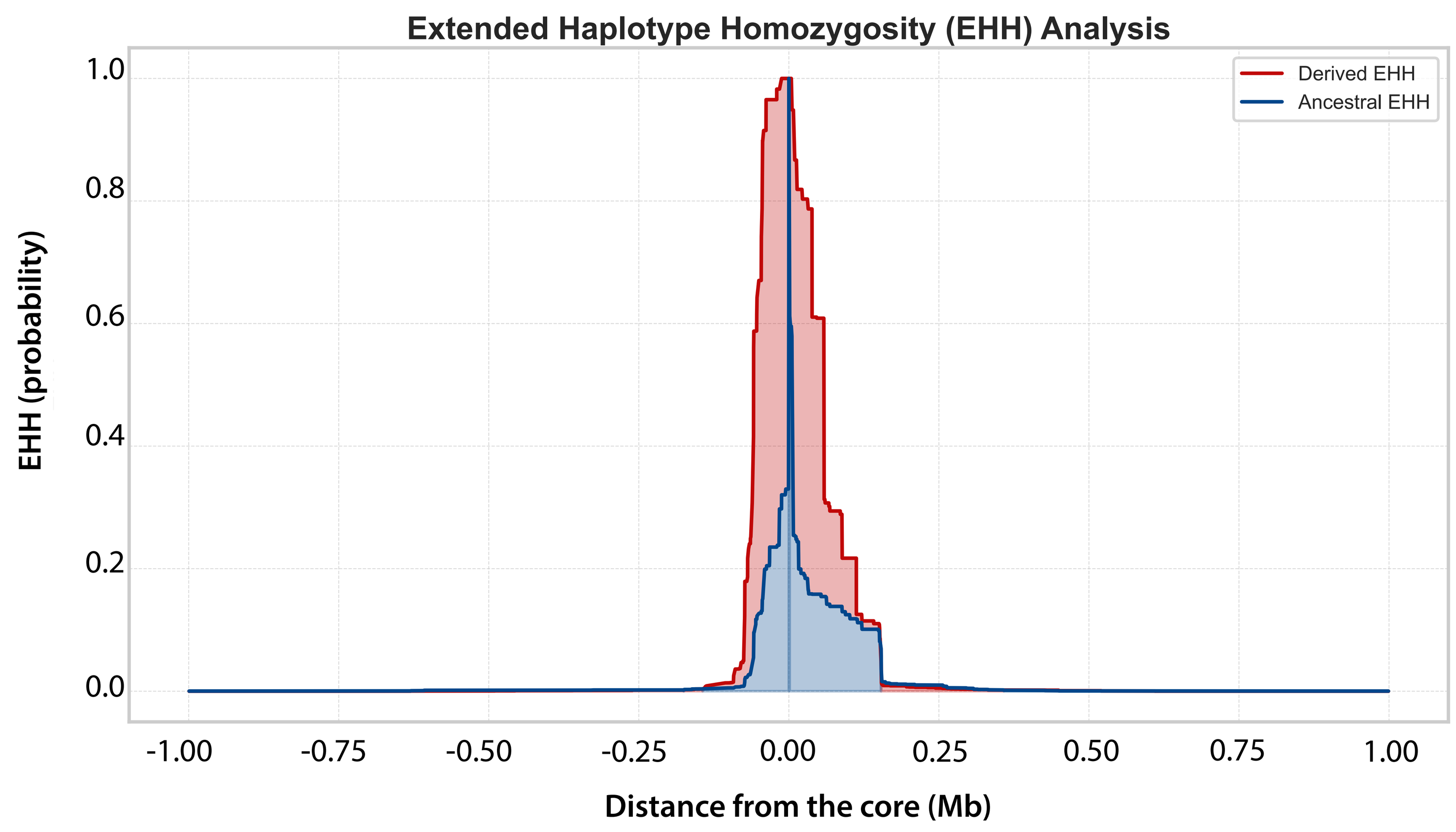


TLR2 (rs11099894)

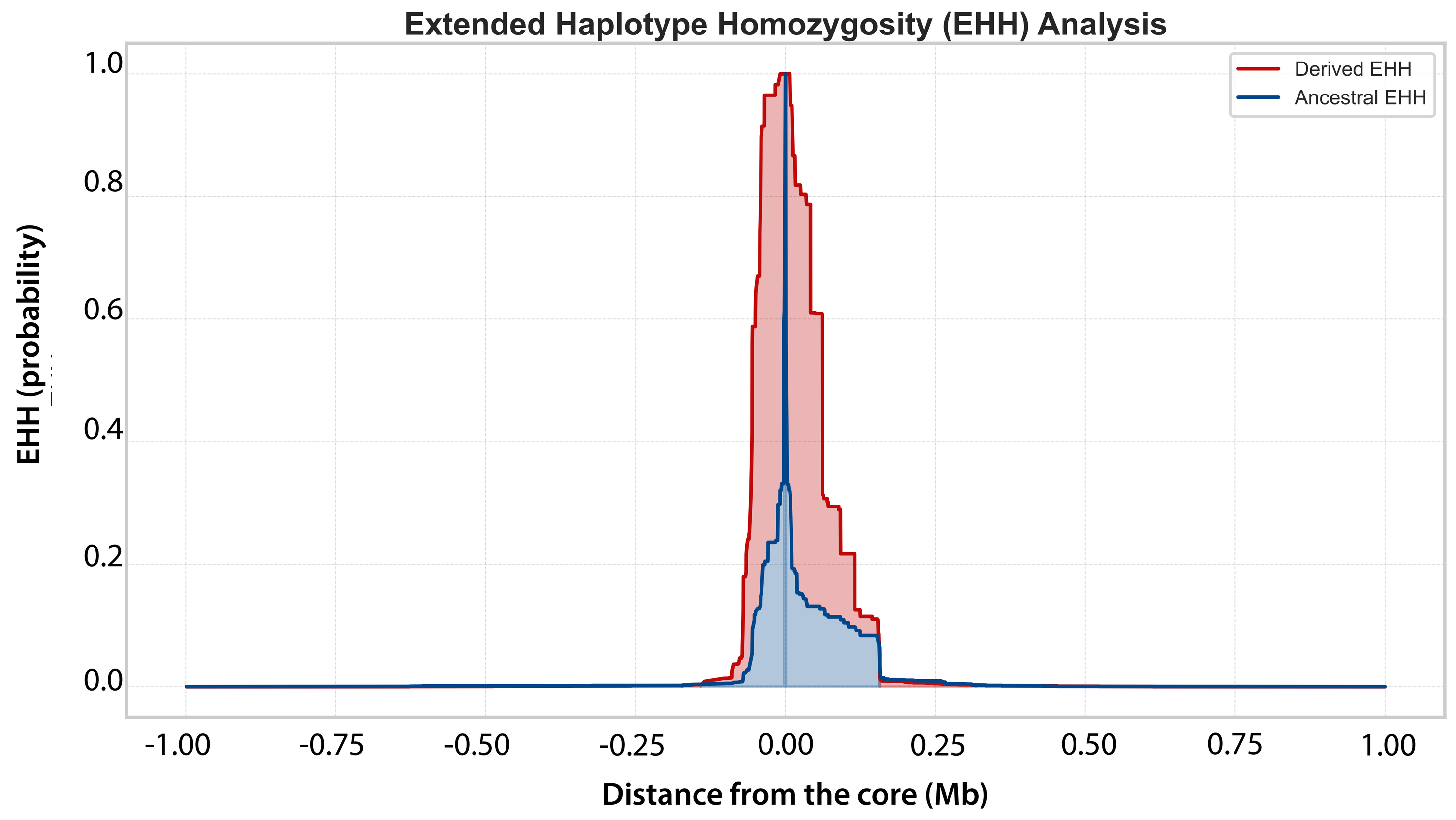


TLR2 (rs13105517)

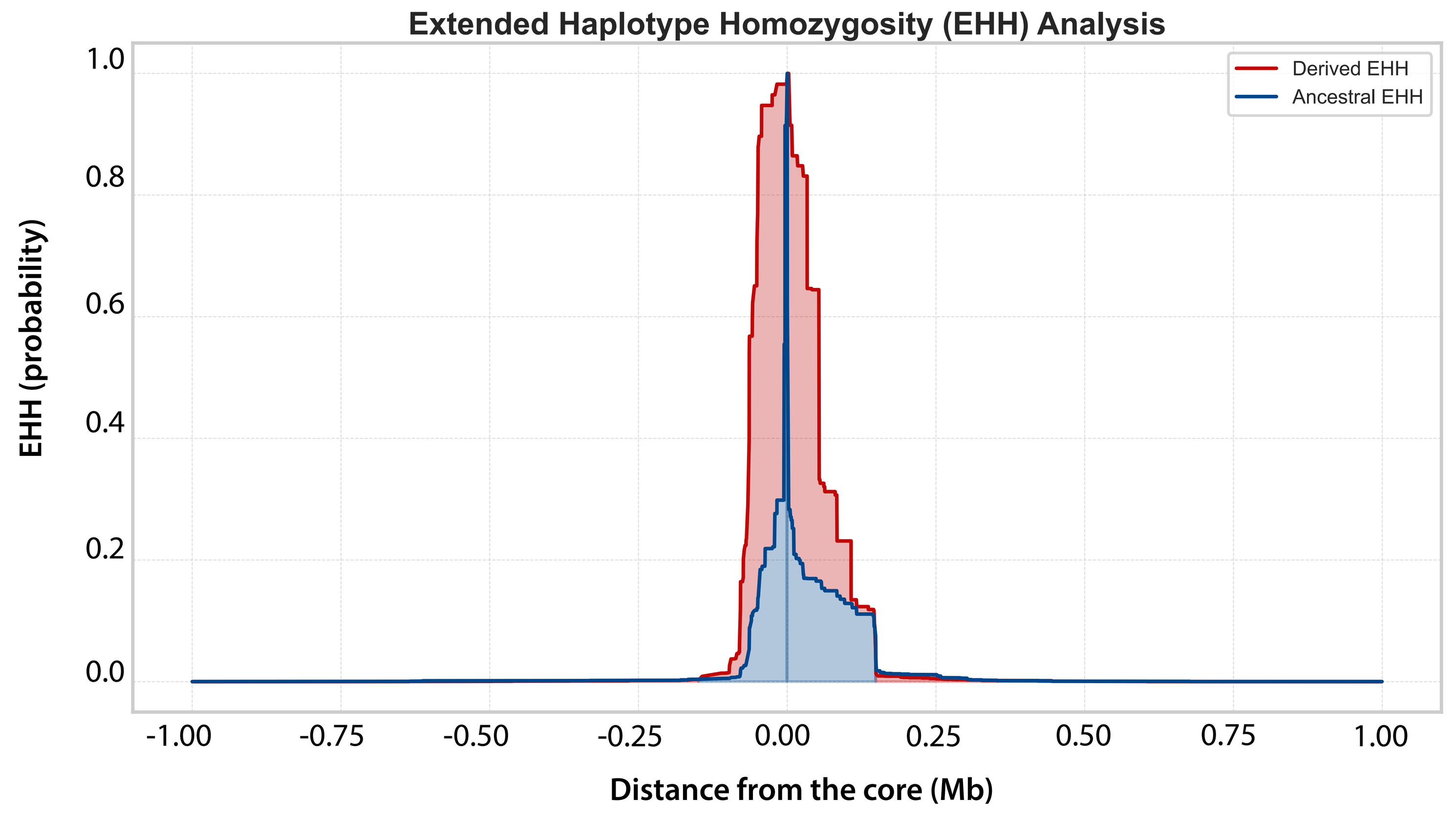


**Figure S6**

LWK

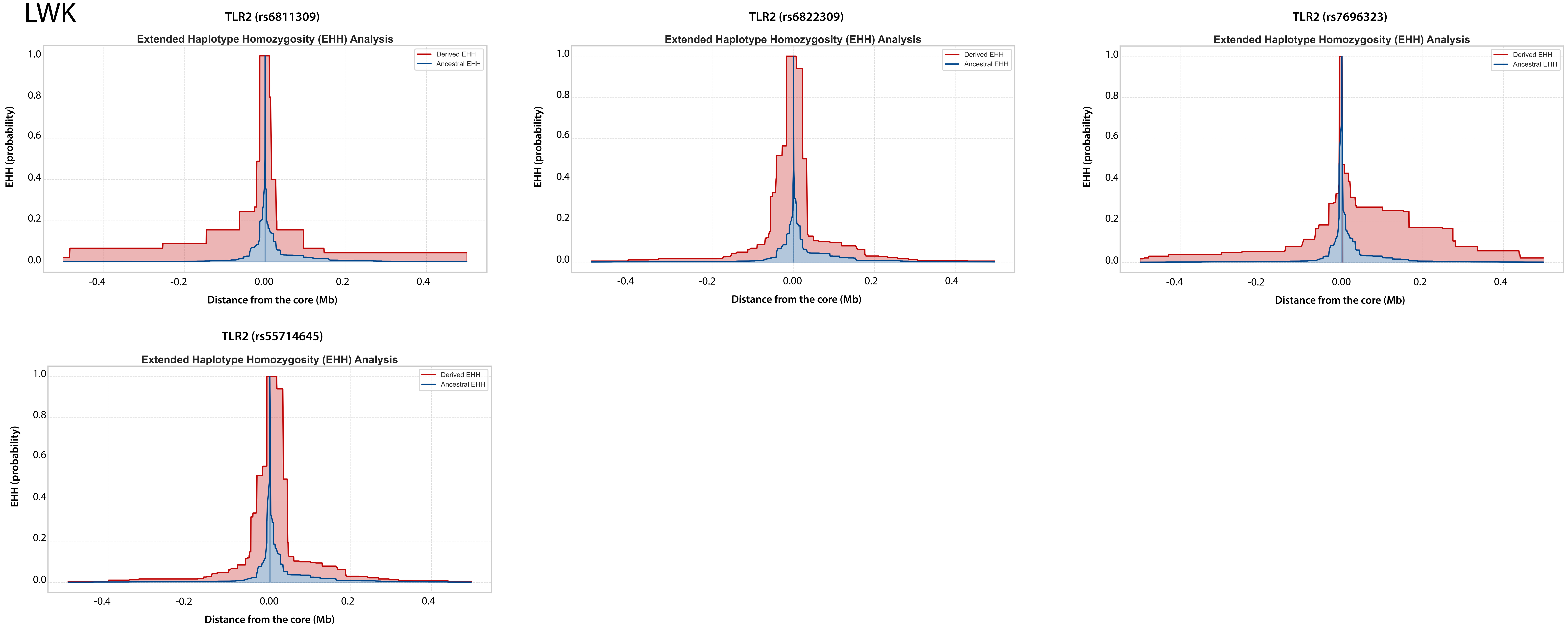


**Figure S6**

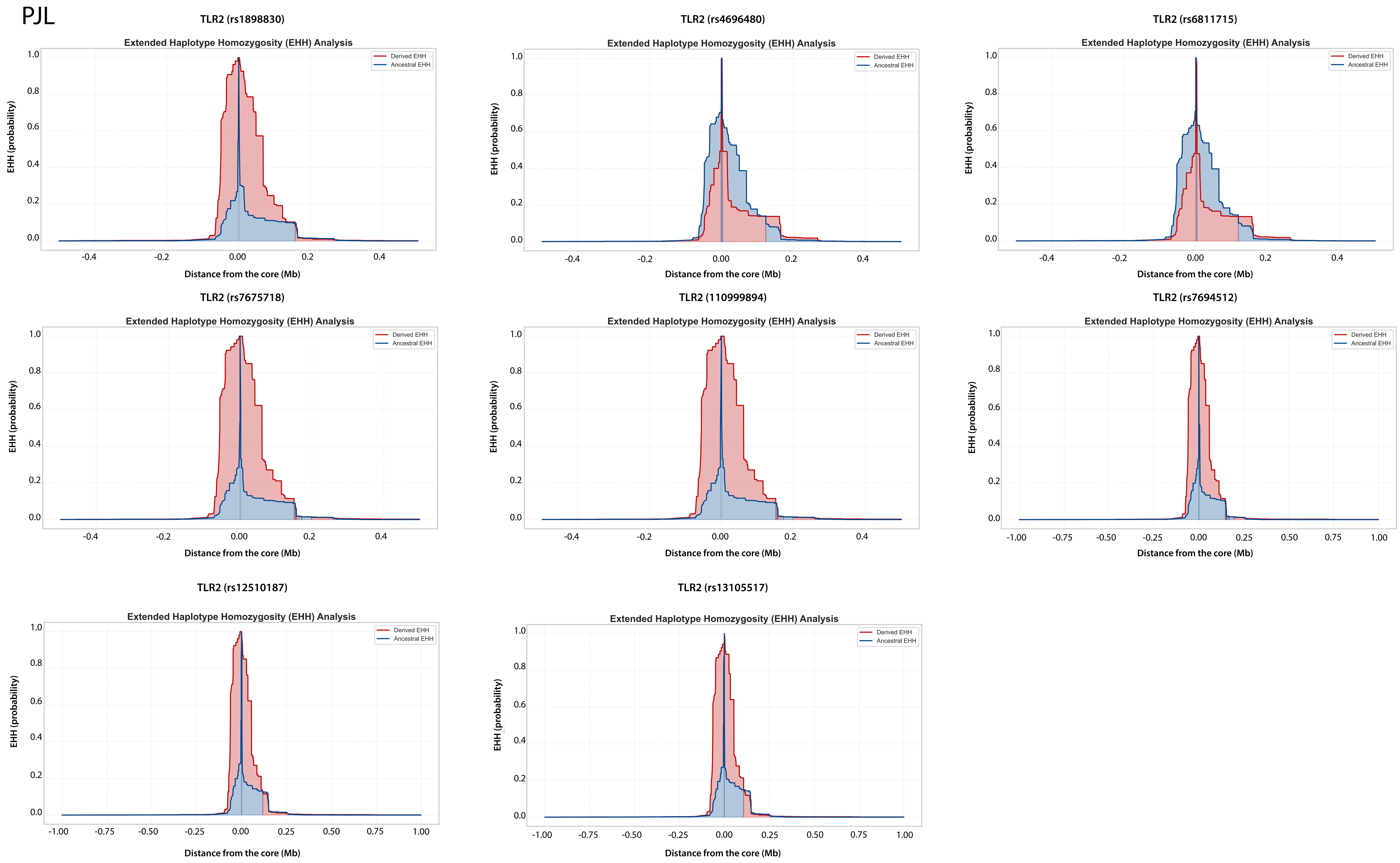
PJL
TLR2 (rs1898830)
TLR2 (rs4696480)
TLR2 (rs6811715)
TLR2 (rs7675718)
TLR2 (110999894)
TLR2 (rs7694512)
TLR2 (rs12510187)
TLR2 (rs13105517)
Extended Haplotype Homozygosity (EHH) Analysis
Derived EHH
Ancestral EHH
EHH (probability)
Distance from the core (Mb)

**Figure S6**

STU

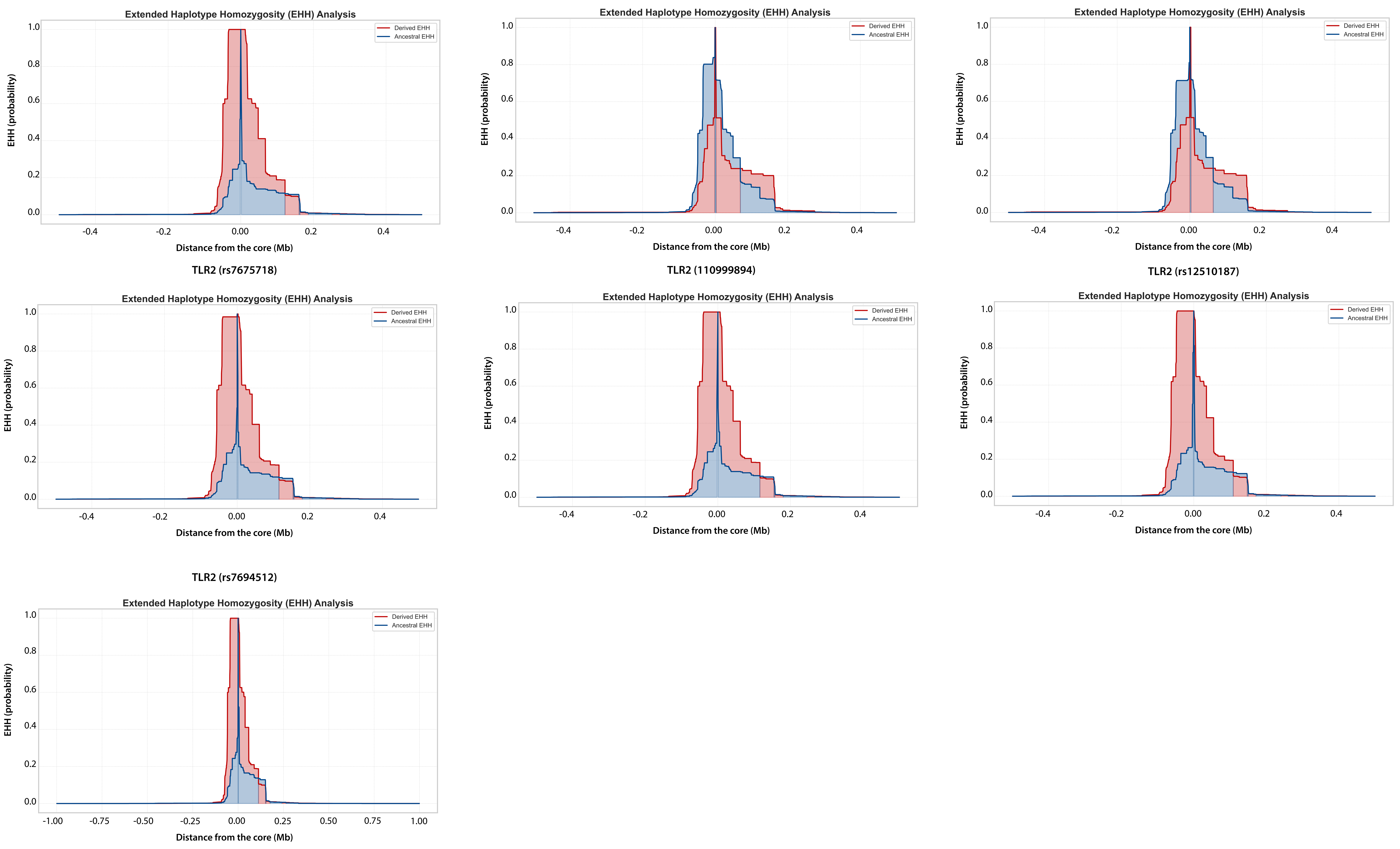


### Figure S6
