## Supplementary material for "The evolutionary landscape of host immunity genes involved in respiratory and other immune-related diseases, and their association with severe COVID-19 outcomes": Figure S10

**Figure S10: Integrated haplotype score (*i*HS) plots for Chromosome 2 in global populations.** Here, we show Manhattan plots of standardized | *i*HS | statistics for single nucleotide polymorphisms (SNPs) on Chromosome 2 in 21 populations from the 1000 Genomes Project. The dashed horizontal lines indicate the threshold for outlier | *i*HS | statistics. We also highlighted the derived alleles at different SNPs across *SLC11A1* with a red dot and list their corresponding rs identifiers near them. The green dots represent ancestral alleles at SNPs across *SLC11A1*, and their corresponding rs identifiers are given next to them.

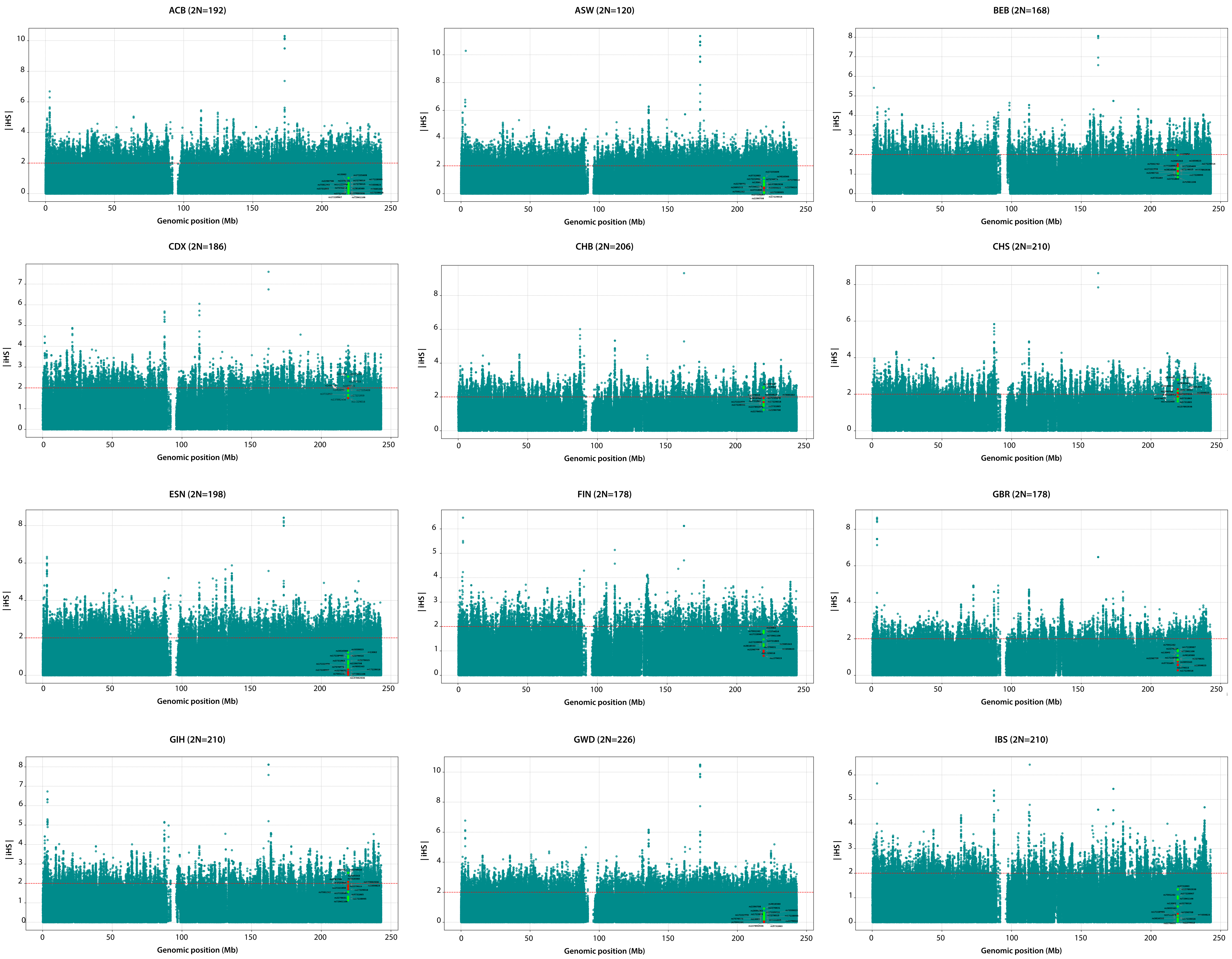
ACB (2N=192)
ASW (2N=120)
BEB (2N=168)
CDX (2N=186)
CHB (2N=206)
CHS (2N=210)
ESN (2N=198)
FIN (2N=178)
GBR (2N=178)
GIH (2N=210)
GWD (2N=226)
IBS (2N=210)
|iHS|
Genomic position (Mb)

### Figure S10

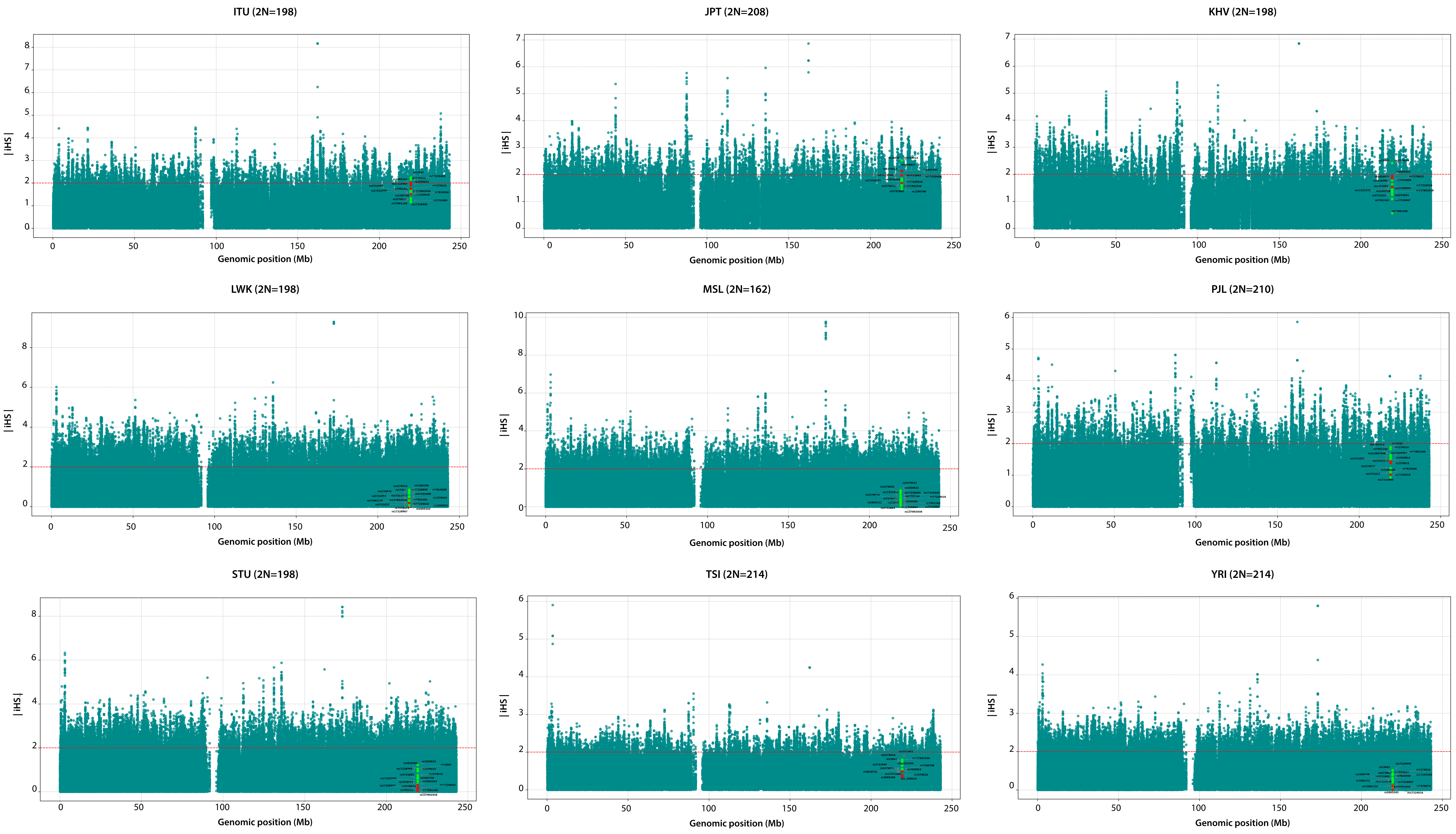
ITU (2N=198)
JPT (2N=208)
KHV (2N=198)
LWK (2N=198)
MSL (2N=162)
PJL (2N=210)
STU (2N=198)
TSI (2N=214)
YRI (2N=214)
|iHS|
Genomic position (Mb)
0
50
100
150
200
250

### Figure S10
