## Supplementary material for "The evolutionary landscape of host immunity genes involved in respiratory and other immune-related diseases, and their association with severe COVID-19 outcomes": Figure S11

**Figure S11: The number of Segregating Sites by Length (*$nS_L$*) plots for Chromosome 2 in global populations.** Here, we show Manhattan plots of standardized | $nS_L$ | statistics for single nucleotide polymorphisms (SNPs) on Chromosome 2 in 21 populations from the 1000 Genomes Project. The dashed horizontal lines indicate the threshold for outlier | $nS_L$ | statistics. We also highlighted the derived alleles at different SNPs across *SLC11A1* with a red dot and list their corresponding rs identifiers near them. The green dots represent ancestral alleles at SNPs across *SLC11A1*, and their corresponding rs identifiers are given next to them.

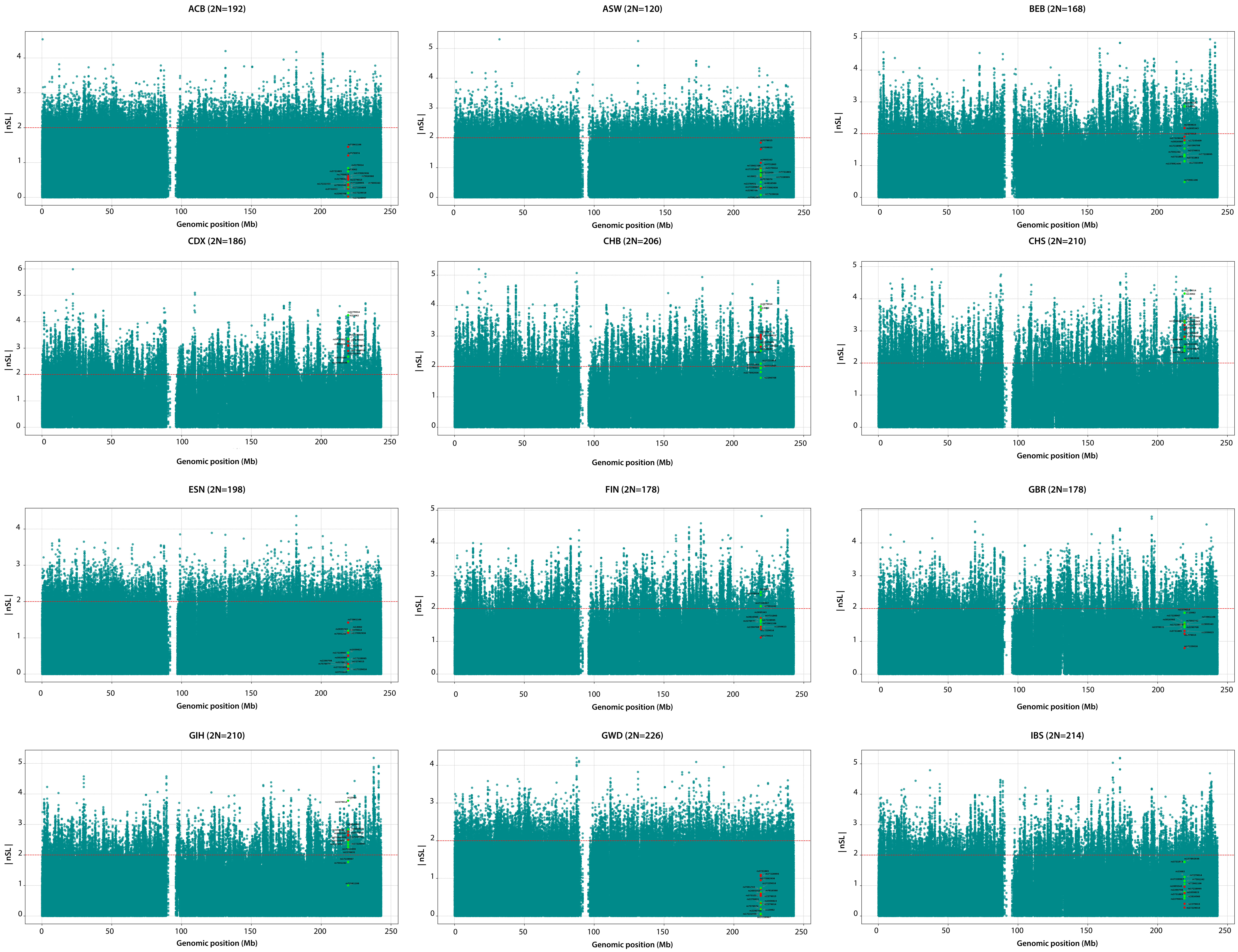
ACB (2N=192)
ASW (2N=120)
BEB (2N=168)
CDX (2N=186)
CHB (2N=206)
CHS (2N=210)
ESN (2N=198)
FIN (2N=178)
GBR (2N=178)
GIH (2N=210)
GWD (2N=226)
IBS (2N=214)
|nSL|
Genomic position (Mb)

Figure S11

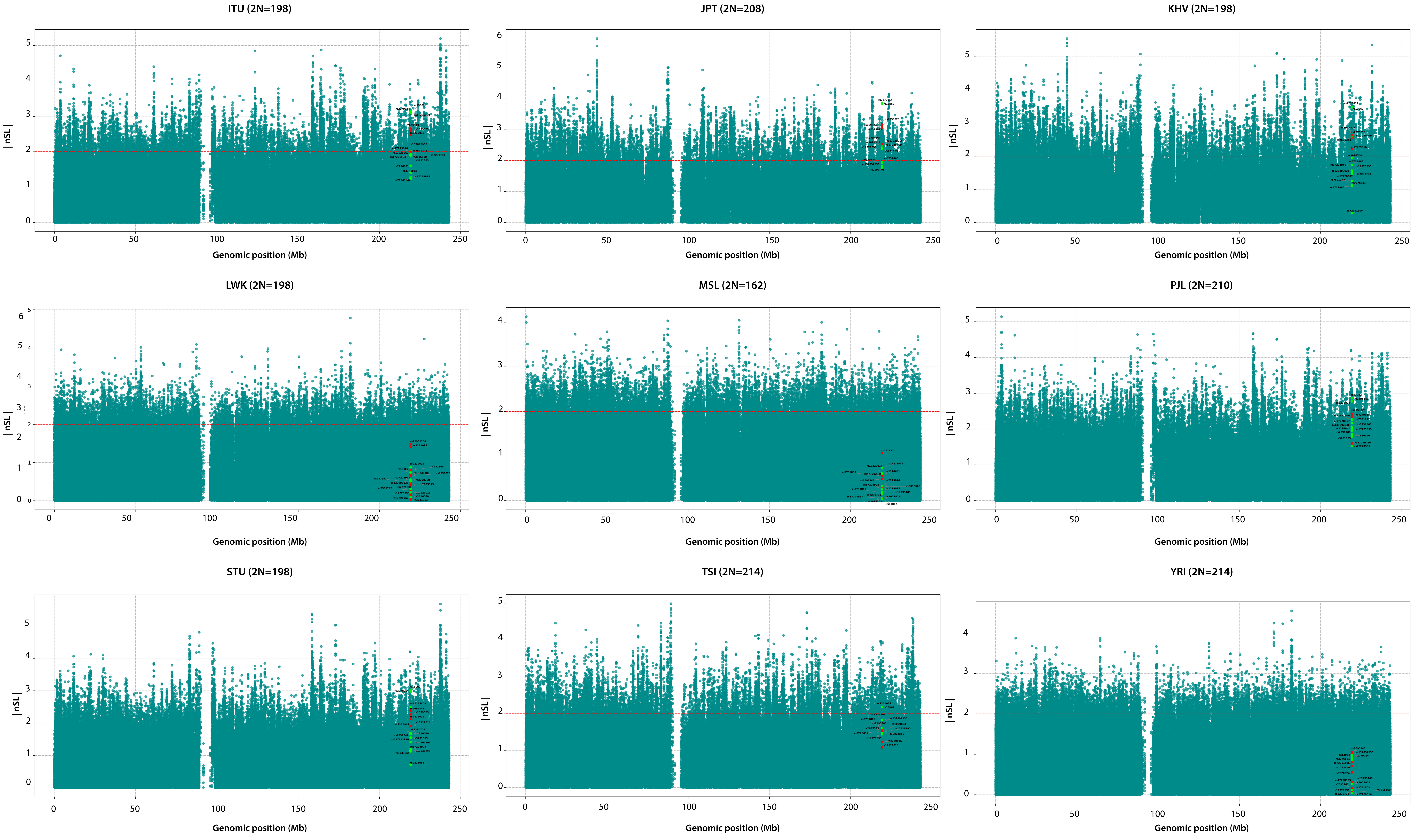
ITU (2N=198)
JPT (2N=208)
KHV (2N=198)
LWK (2N=198)
MSL (2N=162)
PJL (2N=210)
STU (2N=198)
TSI (2N=214)
YRI (2N=214)
|nSL|
Genomic position (Mb)

### Figure S11
